# Apt interpretation of comprehensive lipoprotein data in large-scale epidemiology – disclosure of fundamental structural and metabolic relationships

**DOI:** 10.1101/2021.03.08.21253123

**Authors:** Mika Ala-Korpela, Siyu Zhao, Marjo-Riitta Järvelin, Ville-Petteri Mäkinen, Pauli Ohukainen

## Abstract

**Aims:** Quantitative lipoprotein analytics by NMR spectroscopy is currently commonplace in large-scale studies. One methodology has become widespread and is currently being utilised also in large biobanks. It allows comprehensive characterisation of 14 lipoprotein subclasses, clinical lipids, apolipoprotein A-I and B. The details of these data are conceptualised here in relation to lipoprotein metabolism with particular attention to the fundamental characteristics of subclass particle numbers, lipid concentrations and compositional measures.

**Methods and Results:** The NMR methodology was applied to fasting serum samples from Northern Finland Birth Cohort 1966 and 1986 with 5,651 and 5,605 participants, respectively. All results were highly coherent between the cohorts. Circulating lipid concentrations in a particular lipoprotein subclass arise predominantly as the result of the circulating number of those subclass particles. The spherical lipoprotein particle shape, with a radially oriented surface monolayer, imposes size-dependent biophysical constraints for the lipid composition of individual subclass particles and inherently restricts the accommodation of metabolic changes via compositional modifications. The new finding that the relationship between lipoprotein subclass particle concentrations and the particle size is log-linear reveal that circulating lipoprotein particles are also under rather strict metabolic constraints for both their absolute and relative concentrations.

**Conclusion:** The fundamental structural and metabolic relationships between lipoprotein subclasses elucidated in this study empower detailed interpretation of lipoprotein metabolism. Understanding the intricate details of these extensive data is consequential for the precise interpretation of novel therapeutic opportunities and for fully utilising the potential of forthcoming analyses of genetic and metabolic data in extensive biobanks.

**One-sentence Summary:** NMR spectroscopy facilitates comprehensive characterisation of lipoprotein subclass metabolism and offers additional value to epidemiology, genetics and pharmacology in large-scale studies and biobanks.

**Key Messages:**

- The circulating particle number of a lipoprotein subclass is the defining measure for its lipid concentrations; the particle lipid composition is only in a minor role. The relationship between circulating lipoprotein subclass particle concentrations and the particle size is log-linear.
- The overall structure of lipoprotein subclass particles with a spherical shape and an oriented surface monolayer poses strong size-dependent *biomolecular constraints* for their lipid composition.
- The circulating lipoprotein subclass particle concentrations in humans are *metabolically constraint* for both elemental absolute and relative concentration ranges.
- The smallest HDL particle concentrations are negatively associated with those of large HDL and generally the associations of the smallest HDL particles are similar to those of apolipoprotein B-containing particles.
- The apolipoprotein B-containing particles constitute less than 10% of all lipoprotein particles but carry around two thirds of circulating lipoprotein lipids. LDL and IDL particles amount to almost 90% of all apolipoprotein B-containing particles.
- The supplemental role of lipoprotein subclass data in cardiometabolic risk assessment is slight.
- In the current era of biobanks and big data, the combination of lipoprotein subclass data with drug-target Mendelian randomization analyses provides great scientific synergy, intricate details and potential cost savings in drug development.

**Graphical abstract / key messages:** 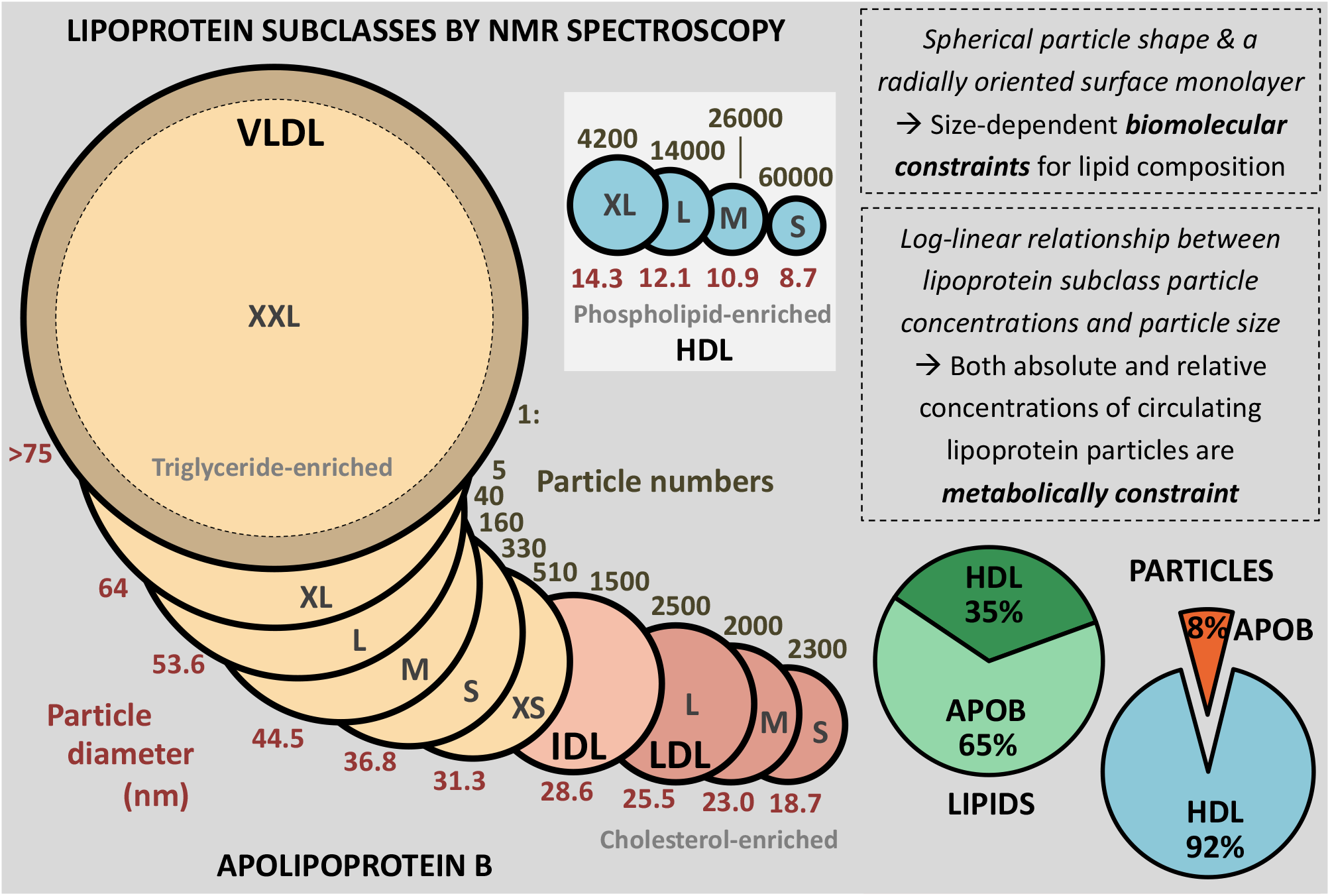

## Introduction

The role of cholesterol in the development of human atherosclerosis has been known for over a hundred years. After the original findings in relation to cholesterol, it took almost 50 years to link cholesterol to circulating low-density lipoprotein (LDL) particles, and reveal the association of LDL cholesterol with coronary heart disease (CHD) in the 1950s.^1^ LDL cholesterol and a few other lipoprotein measures, particularly triglycerides and high-density lipoprotein (HDL) cholesterol, have been in the spotlight of CHD research since then. Thanks to the early discoveries of Brown and Goldstein in the 1970s and 1980s, we now understand the pivotal elements of cholesterol metabolism in atherosclerosis, including efficient pharmacological interventions.^1^

Broader research on the molecular characteristics of circulating lipoprotein particles began after the invention to use sequential preparative ultracentrifugation to physically isolate native lipoprotein particles according to their densities and then further characterise them chemically.^2^ The micellar structure of lipoprotein particles was suggested in 1968 on the basis of nuclear magnetic resonance (NMR) spectroscopy experiments,^3^ leading eventually to the perception of lipoproteins as spherical particles of a non-polar lipid filled core and a surface with radially oriented lipids, together with various apolipoprotein molecules, with polar molecular groups allowing them to be dissolved in a water-based environment, *i*.*e*., blood plasma.^4,5^

Although ultracentrifugation became the workhorse in lipoprotein analytics, the tediousness of the process, particularly with a tight density scale, prevented large studies with high resolution of lipoprotein fractions.^4^ However, in the 1990s NMR spectroscopy was introduced as a cost-effective methodology to acquire detailed large-scale data on lipoprotein subclasses. A few methodologies were developed with slightly different definitions and numbers of lipoprotein subclasses.^6,7^ The methodology invented and developed in our research team^6^ was extended to a metabolomics platform to quantify over 200 metabolic measures, including comprehensive characterisation of 14 lipoprotein subclasses. Over the last 10 years this particular platform has become common in large-scale epidemiology and genetics,^8–10^ and it is currently been applied to analyse all the 500,000 serum samples in the UK Biobank, with over 100,000 samples already analysed.^11^ Some 300 publications with applications of this methodology are currently available and more are written with an increasing speed; when the extensive and unique lipoprotein data will be publicly available in the UK Biobank, the number of publications is likely to surge.

Almost all of epidemiological and genetic applications of the lipoprotein data have focused on concentration measures only, even though the compositional data are automatically incorporated in the analytical results for each serum sample.^9,10^ One reason for the reluctance of researchers to include the compositional data may be its unfamiliarity both in epidemiology and clinical chemistry, not to mention clinical studies. In addition, the concentration data for 14 lipoprotein subclasses consist already of 98 measures (plus apolipoproteins B and A-I as well as various standard lipoprotein lipid measures), thus adding 70 compositional measures considerably complicates statistical analyses and presentation of results. Nevertheless, recent applications have indicated that detailed understanding of the treatment effects of novel lipid-modifying drug targets would remarkably benefit from high-resolution comprehensive molecular data on lipoprotein subclasses and that compositional data may have an independent role in the risk assessment and understanding of lipoprotein metabolism.^12–15^ It is therefore pertinent to discuss the comprehensive lipoprotein subclass data of this particular NMR platform in detail and specify the fundamental characteristics of the concentration and compositional data and their relationships. This is done for the first time in this work.

## Methods

### Lipoprotein subclasses by NMR spectroscopy

The separation of lipoprotein subclasses in proton NMR spectroscopy is based on particle size. In more detail, the anisotropy of the magnetic susceptibility, due to the radially oriented surface monolayer of the spherical particles, inherently leads to a size-dependent frequency shift for the NMR resonances of lipoprotein lipids.^16^ The NMR data in this particular platform are calibrated via high-performance liquid chromatography with the platform resolution being 14 lipoprotein subclasses, defined by their particle size as follows: six very-low-density lipoprotein (VLDL) particle categories; extremely large (XXL-VLDL, with average particle diameter >75 nm), very large (XL-VLDL, 64 nm), large (L-VLDL, 53.6 nm), medium (M-VLDL, 44.5 nm), small (S-VLDL, 36.8 nm), and very small (XS-VLDL, 31.3 nm); intermediate-density lipoprotein particles (IDL, 28.6 nm), large (L-LDL, 25.5 nm), medium (M-LDL, 23.0 nm), and small LDL particles (S-LDL, 18.7 nm); and very large (XL-HDL, 14.3 nm), large (L-HDL, 12.1 nm), medium (M-HDL, 10.9 nm) and small HDL particles (S-HDL, 8.7 nm).^9,17^ Independent verification for the robust NMR resolution with respect to the number of lipoprotein subclasses was published by Mihaleva and co-workers with an in-depth handling of the statistical grounds.^18^ Particle, triglyceride, phospholipid, cholesteryl ester and free cholesterol concentrations are quantified for all 14 subclasses, allowing also calculation of the relative lipid contents (*i*.*e*., lipid composition) for each lipoprotein subclass. The platform also provides all key traditional lipid measures (*e*.*g*., total triglycerides, total cholesterol and HDL cholesterol) as well as apolipoprotein B and A-I concentrations. This NMR metabolomics platform has been widely used in epidemiology and genetic studies over the last 10 years^9–11,17^ and the general methodological issues have been published and discussed previously.^8,17,19^

### Population Cohorts

The Northern Finland Birth Cohort 1966 (NFBC66) with data for 5,651 participants (at 46 years of age) were studied as the primary cohort and the NFBC86 with data for 5,605 participants (at 16 years of age) as a replication cohort. More details on the NFBCs can be found in the Supplementary Note. The selection or the traits of the cohorts are not consequential in this study focusing on overall lipoprotein subclass particle characteristics and associations as the existing literature already affirms them highly coherent in and between various large-scale studies.^9,10,12,13,17,20–24^

### Statistical analyses and concepts

The lipoprotein subclass information can be partitioned into three key categories of variables: 1) *circulating particle concentrations* (the particle numbers are within the range of 0.1 nmol/L and 10 µmol/L), 2) *circulating lipid concentrations* (within the range of 0.1 and 10 mmol/L) and 3) *particle lipid compositions* as the percentage of a certain lipid class of total lipids (mol%) per a subclass particle. Apolipoprotein B is the key protein component in VLDL, IDL and LDL particles and they are often called apolipoprotein B-containing particles. HDL particles do not contain apolipoprotein B but are abundant in apolipoprotein A-I. All lipoprotein particles contain also other apolipoprotein molecules, but they cannot be quantified by NMR.^25^ Spearman’s rank correlations (adjusted for sex) were used in all correlation analyses in this work. Twenty-four principal components explained >99% of variation in the 174 lipoprotein measures (98 concentrations and 70 compositions for the 14 lipoprotein subclasses, 4 traditional lipid measures and apolipoprotein A-I and B). A multiple comparison Bonferroni-corrected P-value threshold of 0.002 (i.e., 0.05/24) was thus used to denote evidence in favour of an association.^26^ All statistical analyses were performed in the R statistical platform (version 3.6.2).

## Results

### Characteristics of lipoprotein subclass particles and their concentration attributes

Figure 1. illustrates the 14 lipoprotein subclasses and their relative lipid content (see **Figure S1** for replication in the NFBC86). The compositional data are also visualised in the upper part of **Figure 2** in comparison to the concentration data (identical data are shown for the NFBC86 in **Figure S2**). The VLDL subclasses are the most triglyceride-enriched with variation from 69% to 17% of total lipids from XXL-VLDL to XS-VLDL. The IDL and LDL particles are all rich in cholesteryl esters (around 50%) as well as free cholesterol (around 20%). All HDL particles are enriched in phospholipids (around 50%) and very poor in triglycerides.

**Figure 1.**
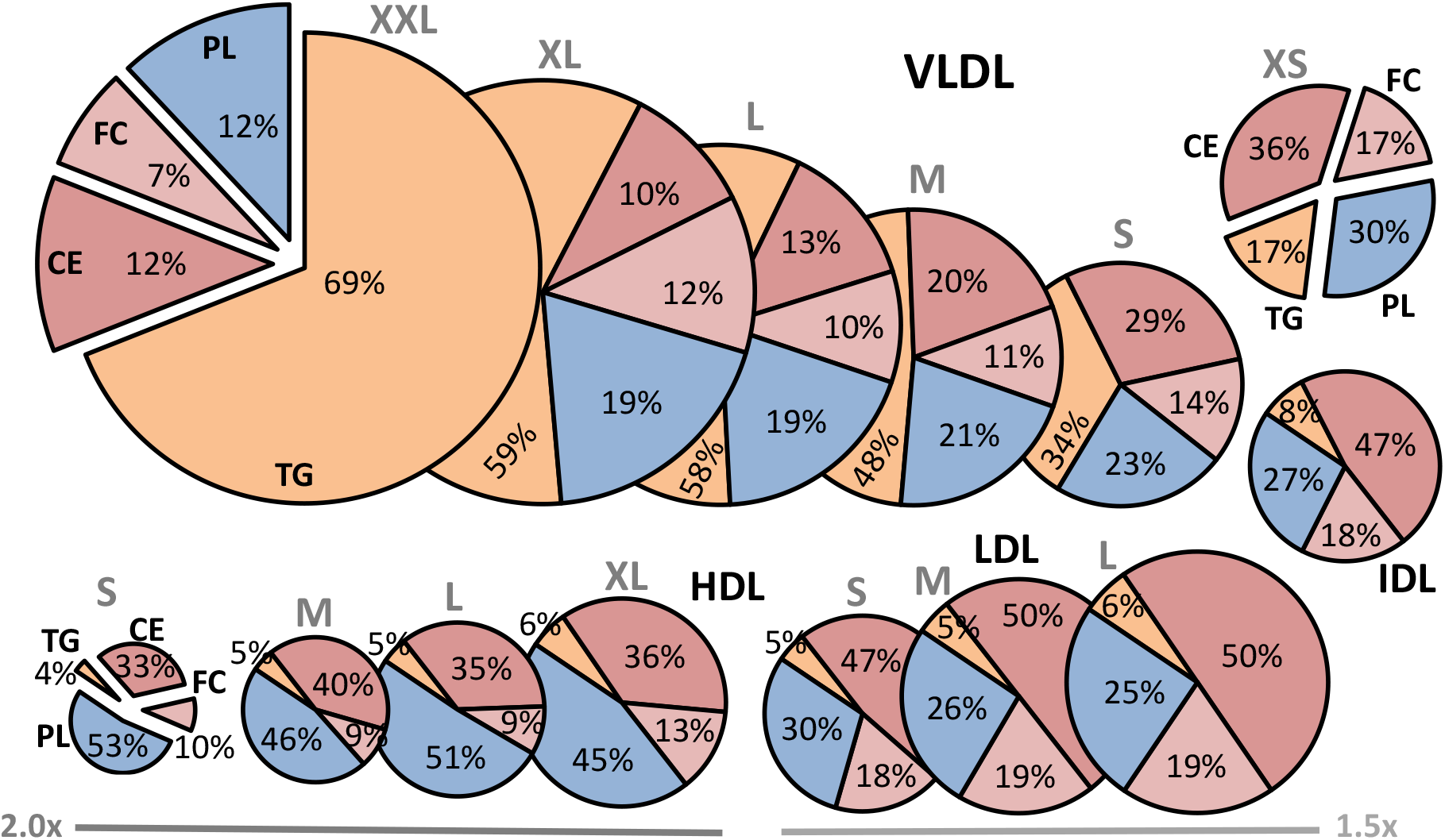
The relative lipid compositions of the 14 lipoprotein subclasses.The NMR platform resolution is 14 lipoprotein subclasses, defined by their particle size as follows: six VLDL particle categories; XXL-VLDL (with average particle diameter >75 nm), XL-VLDL (64 nm), L-VLDL (53.6 nm), M-VLDL (44.5 nm), S-VLDL (36.8 nm), and XS-VLDL (31.3 nm); IDL (28.6 nm), L-LDL (25.5 nm), M-LDL (23.0 nm), and S-LDL (18.7 nm); and XL-HDL (14.3 nm), L-HDL (12.1 nm), M-HDL (10.9 nm) and S-HDL (8.7 nm).^9,17,19^ Note that the size of LDL and HDL particles in the figure is multiplied by 1.5 and 2.0, respectively. The data are mean values for 5,651 participants in NFBC66. VLDL, very-low-density lipoprotein; IDL, intermediate-density lipoprotein; LDL, low-density lipoprotein; HDL, high-density lipoprotein; XXL, extremely large; XL, very large; L, large; M, medium; S, small; XS, very small; TG, triglycerides; PL, phospholipids; CE, cholesteryl esters; FC, free cholesterol.

**Figure 2.**
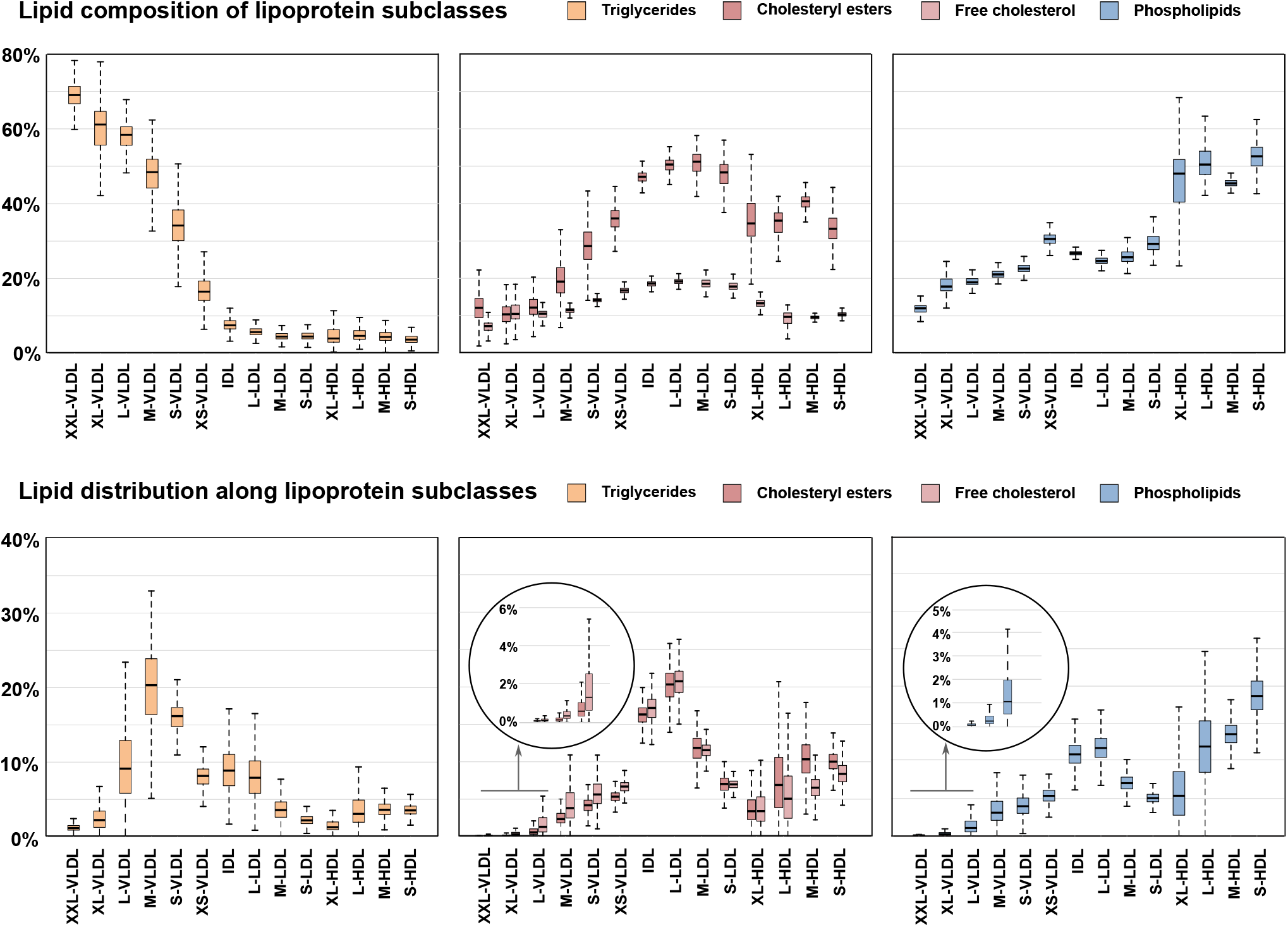
The circulatory and compositional characteristics of lipoprotein subclasses.The lipid distribution along the lipoprotein subclasses illustrates how a certain lipid class is distributed in the blood stream among all the lipoprotein subclasses (i.e., the sum of all percentages in each inset is 100%). The composition of lipoprotein subclasses depicts what are the relative lipid contents in each subclass particle category (i.e., the sum of the percentages of all the lipid classes for each subclass is 100%). The data are from the NFBC66 including 5,651 participants; each box plot shows the median within the interquartile range (IQR) and the minimum (Q1-1.5*IQR) and maximum (Q3 + 1.5*IQR) values with potential outliers. The abbreviations are as explained in the caption for Figure 1.

Over 50% of circulating triglycerides are transported in VLDL particles, some 20% in IDL and LDL, and less than 20% in HDL particles (lower part of **Figure 2** and **S2**). Roughly 50% of circulating cholesterol (both free and esterified) travels in IDL and LDL particles and some 20-30% in HDL particles. Around 50% of lipoprotein phospholipids in the circulation are in HDL particles. The size-dependent opposite compositional behaviour of triglycerides and cholesteryl esters in the apolipoprotein B-containing subclass particles is pronounced.

The circulating subclass lipid concentrations depend on both, the lipid composition of a subclass particle and the concentration (*i*.*e*., the number) of those particles in the blood stream. The distributions for subclass particle concentrations are shown in **Figure 3** (and in **Figure S3** for the NFBC86). The HDL particles are by far the most abundant with concentrations in a micromolar range, the IDL and LDL particles being at the range of 0.1-0.3 micromolar and the largest VLDL particles at the range of 0.1 – 10 nanomolar. Thus, on average, there are some 60,000 S-HDL and 1,500 IDL particles per one XXL-VLDL particle in the circulation; 92% of all lipoprotein particles are HDL particles (**Figure 3**). Therefore, although the volume of an XXL-VLDL particle is approximately 640 times that of an S-HDL particle (and 18 times that of an M-LDL particle), the variation in the particle lipid compositions within a subclass is clearly minor with respect to the variation in corresponding particle concentrations. The circulating particle concentrations are therefore the defining measures for the lipoprotein subclass lipid concentrations. These particle concentrations are log-linearly associated with the subclass particle size as illustrated in **Figure 4**. This intriguing finding denotes that numbers of circulating lipoprotein particles are fundamentally restricted, both in a relative manner and in absolute concentrations (*e*.*g*., the number of VLDL particles must be appropriate for the circulating LDL cholesterol to remain within physiologically relevant limits).

**Figure 3.**
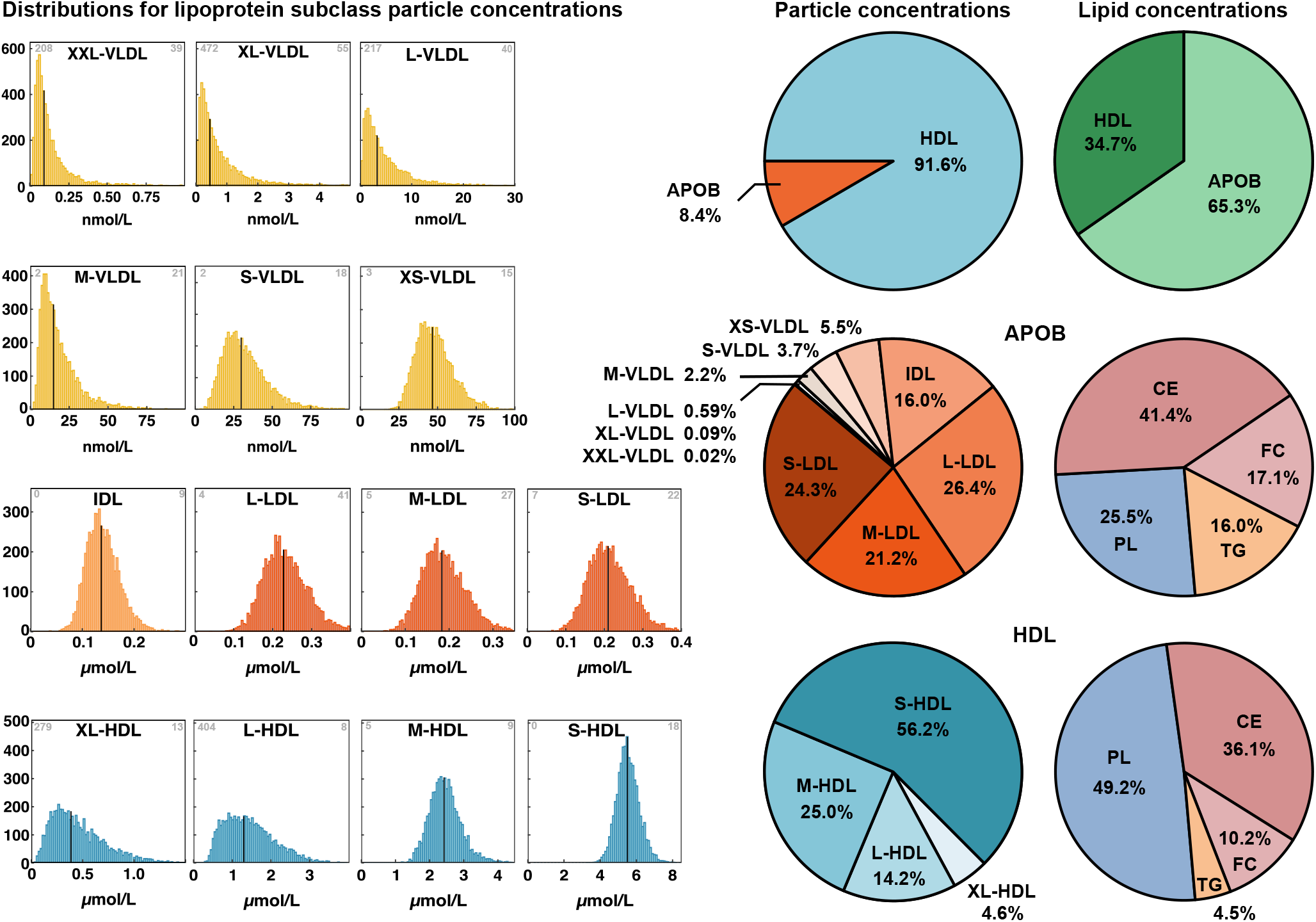
The distributions of particle concentrations for each lipoprotein subclass.The data are from the NFBC66 including 5,651 participants. The grey number in the upper-left corner identifies the number of samples for which the particle concentration is zero and in the upper-right corner for how many high concentration values were cut off from the drawn distribution. Note that the concentration scale is nmol for the VLDL, IDL and LDL particles and µmol for the HDL particles. The black vertical lines denote the median concentration values. Various proportions of lipoprotein particles and lipids are shown in the pie charts as mean values for the 5,651 participants in NFBC66. The abbreviations are as explained in the caption for Figure 1.

**Figure 4.**
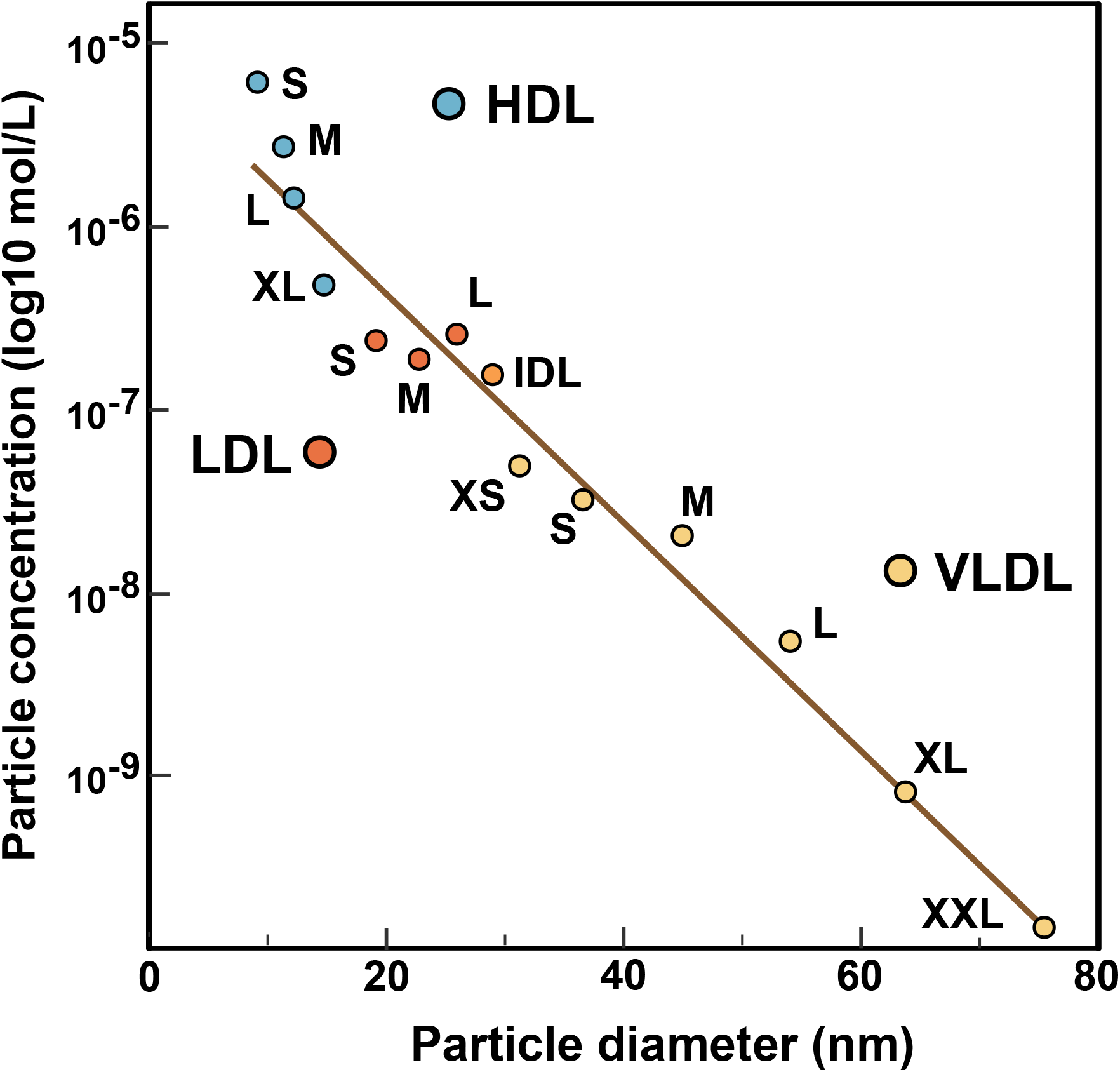
The log-linear relationship between the circulating lipoprotein subclass particle concentration and the particle diameter.The data and abbreviations are as explained in the caption for Figure 1.

The overall outlook on lipoprotein lipid transport, due to large differences in the subclass particle transport volumes, is quite different from the particle view; 65% of all lipoprotein lipids in the circulation are transported in apolipoprotein B-containing particles (**Figure 3**). On average LDL particles amount to over 70% and the three largest VLDL subclasses only less than a percent of circulating apolipoprotein B-containing particles. The smallest HDL particles constitute over 50% of all circulating HDL particles. The apolipoprotein B-containing particles are most abundant in cholesteryl esters (over 40 mol% of all lipids) and the HDL particles in phospholipids (almost 50 mol%) (**Figure 3**). The absolute concentrations (mmol/L) for total lipids and the main lipids in all the 14 lipoprotein subclasses are shown in **Figure 5 (**and in **Figure S5** for the NFBC86) together with a summary comparison between apolipoprotein B-containing particles and HDL particles.

**Figure 5.**
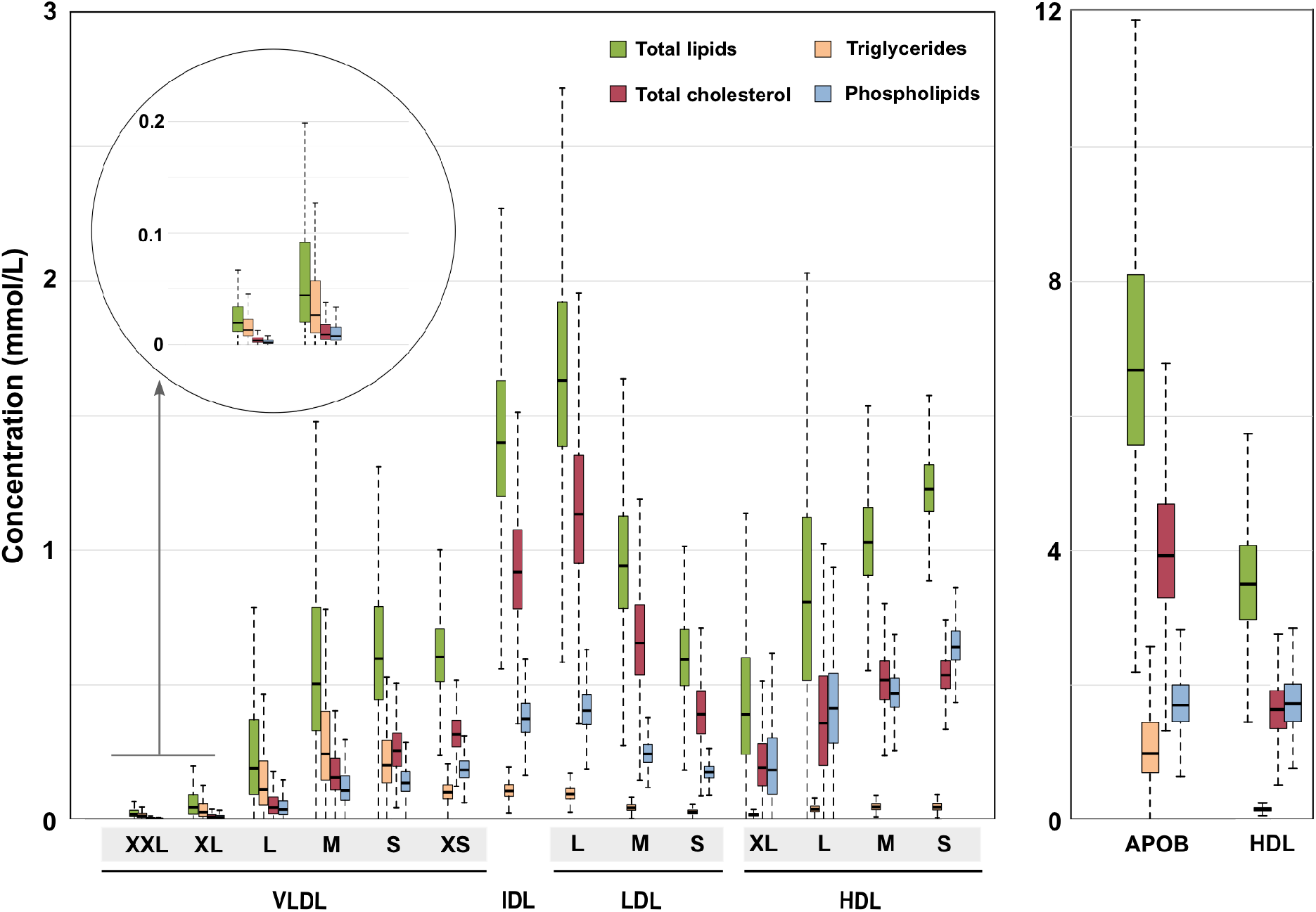
The absolute circulating lipid concentrations for each lipoprotein subclass and the corresponding summary measures for the apolipoprotein B-containing lipoprotein particles and HDL particles.The data and abbreviations are as explained in the captions for Figure 1 and 2.

### Associations between standard lipid and apolipoprotein measures

The standard lipid measures (also based on the NMR platform) behave as expected as illustrated in **Figure 6A** (and in **Figure S6A** for the NFBC86 data).^10^ The associations between apolipoprotein B and LDL cholesterol, total cholesterol and triglycerides are positive and strong. The negative association between HDL cholesterol and triglycerides is compelling. Apolipoprotein A-I is strongly positively associated with HDL cholesterol. Also, as expected, associations between apolipoprotein A-I and HDL particle concentrations and apolipoprotein B and the corresponding particle concentrations are strong (**Figure S7**). On the other hand, associations between apolipoprotein A-I and B concentrations (g/L) as well as between HDL particle and apolipoprotein B-containing lipoprotein particle concentrations (mol/L) are very weak. This is explained by negative associations between apolipoprotein B and large and medium HDL particles and a counterbalancing effect due to a positive association between apolipoprotein B and the smallest HDL particles (**Figure S8**).

**Figure 6.**
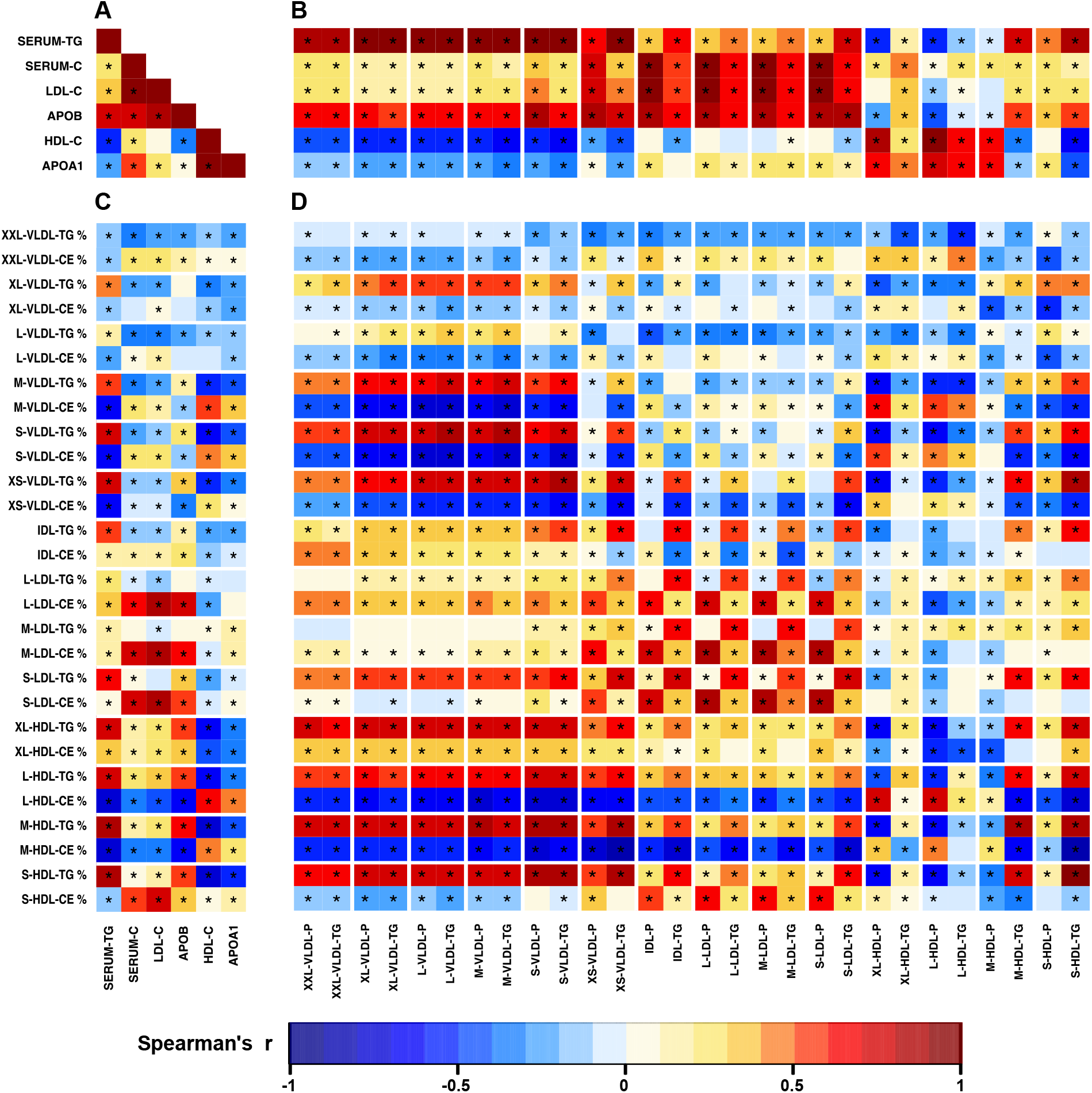
The associations between traditional lipoprotein lipid measures (A), between traditional lipoprotein lipid measures and lipoprotein subclass particle and triglyceride concentrations (B), between traditional lipoprotein lipid measures and lipoprotein subclass lipid composition for triglycerides and cholesteryl esters (C), and between lipoprotein subclass particle and triglyceride concentrations and lipoprotein subclass lipid composition for triglycerides and cholesteryl esters (D). The colour key refers to Spearman’s rank correlation coefficients the NFBC66 including 5,651 participants (adjusted for sex). Twenty-three principal components explained >99% of variation in the 174 lipoprotein measures (98 concentrations and 70 compositions for the lipoprotein subclasses, 4 traditional lipid measures and apolipoprotein A-I and B). Thus, a P-value threshold of 0.002 (*i*.*e*., 0.05/24) was used to denote evidence in favour of an association (marked *in the maps). The %-sign refers to the compositional measures (i.e., the percentage of a lipid class concentration of the total lipid concentration in a particular lipoprotein subclass). Serum-TG, total circulating triglyceride concentration; Serum-C, total circulating cholesterol concentration; LDL-C, LDL cholesterol; APOB, apolipoprotein B; HDL-C, HDL cholesterol; APOA-I, apolipoprotein A-I. Other abbreviations are as explained in the caption for Figure 1.

### Associations between standard lipid measures and lipoprotein subclass concentrations

The associations between the 14 lipoprotein subclass measures and the standard lipid measures are all logical (**Figure 6B** and **S6B**). The associations of apolipoprotein B with all VLDL, IDL and LDL subclass concentration measures are positive and strong. VLDL subclass concentration measures associate positively with serum triglycerides and negatively with HDL cholesterol. Associations between LDL cholesterol and IDL and LDL subclass concentration measures are positive and strong. The associations with HDL subclass measures are heterogeneous, however, HDL cholesterol and apolipoprotein A-I correlate positively with all HDL subclass particle concentrations.

### Associations between standard lipid measures and lipoprotein subclass compositions

The triglyceride composition of all lipoprotein subclasses is positively associated with circulating triglyceride concentration, except in the case of XXL-VLDL **(Figure 6C** and **S6C**). The cholesteryl ester composition of all LDL subclass particles is strongly positively associated with circulating total and LDL cholesterol as well as apolipoprotein B concentrations. Circulating HDL cholesterol and apolipoprotein A-I concentrations are negatively associated with the triglyceride composition of all VLDL and IDL particles, positively associated with the cholesteryl ester composition of S-, M, and L-HDL particles, but the association is negative for XL-HDL particles.

### Associations between lipoprotein subclass concentrations and compositions

An opposite compositional behaviour is typical between triglycerides and cholesteryl esters **(Figure 6D** and **S6D**). For example, high circulating particle and triglyceride concentrations for apolipoprotein B-containing lipoprotein particles associate with triglyceride enrichment and diminished composition of cholesteryl esters in M- and L-HDL particles. Similar strong relationships exist between the concentrations as well as triglyceride and cholesteryl ester compositions of XS-, S-, M-, L-, and XL-VLDL particles. The triglyceride composition of all HDL subclass particles is consistently positively associated with all apolipoprotein B-containing lipoprotein particle and triglyceride concentrations. The circulating concentrations of M-, L-, and XL-HDL particles are negatively associated with their triglyceride compositions.

### Associations between lipoprotein subclass concentrations

The associations between all the apolipoprotein B-containing lipoprotein subclass concentration measures form a coherent pattern and reflect the continuous metabolism from VLDL to LDL particles in the blood stream (**Figure 7A** and **S9A**). Positive associations between adjacent subclasses are high and there is a general trend of weakening positive associations along the line of the metabolic direction from large VLDL to LDL particles. The association pattern appears different for XL- and L-HDL in comparison to M- and S-HDL subclasses. The subclass particle concentrations of large and medium HDL subclasses associate negatively with all VLDL subclasses but the associations for the smallest HDL subclass are in the opposite direction. Similarly, S-HDL particle concentrations are negatively associated with those of XL- and L-HDL. The circulating triglyceride concentration in every apolipoprotein B-containing subclass associates consistently positively with the corresponding measure in both M- and S-HDL.

**Figure 7.**
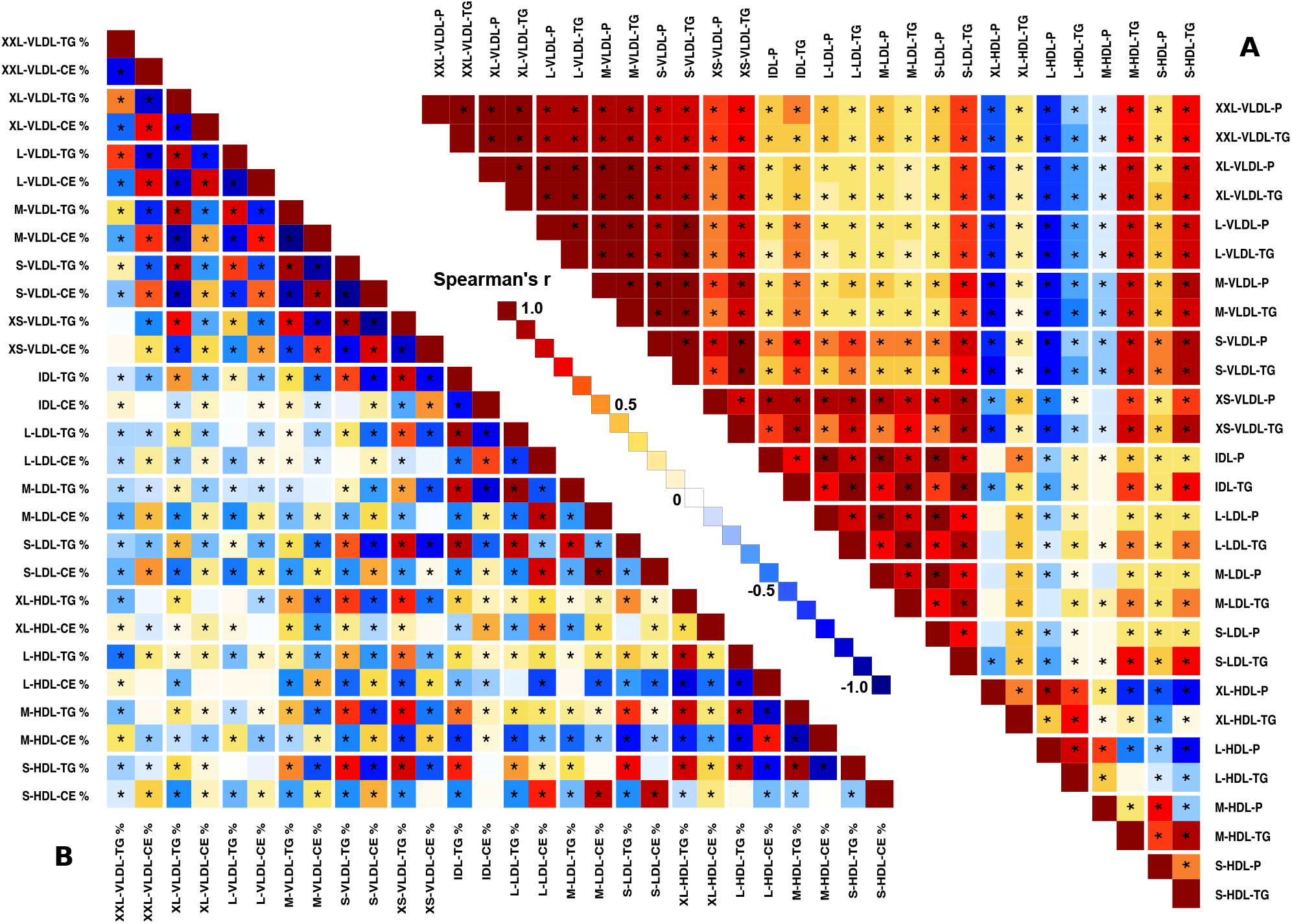
The associations between lipoprotein subclass particle and triglyceride concentrations (A) and between lipoprotein subclass lipid composition for triglycerides and cholesteryl esters (B).The data and abbreviations are as explained in the caption for Figure 6.

### Associations between lipoprotein subclass compositions

The associations between lipoprotein subclass lipid compositions **(Figure 7B** and **S9B**) are generally a lot weaker and more heterogeneous than the corresponding associations between the concentration measures (**Figure 7A** and **S9A**). A global feature is that the triglyceride compositions of almost all lipoprotein subclass particles are strongly positively associated. These compositional triglyceride associations are strong in each major fraction of particles, *i*.*e*., within VLDL, IDL and LDL as well as HDL, but also mostly between these fractions with only a few triglyceride compositions in the largest VLDL subclasses showing either very weak or negative associations. Overall triglyceride and cholesteryl ester compositions tend to associate negatively, particularly within the major fractions of particles.

A correlation heatmap between all the lipoprotein subclass concentration, composition and standard lipid measures as well as apolipoprotein B and A-I are shown in **Figure S10** for the NFBC66 and in **Figure S11** for the NFBC86. The numerical data for these correlations are available in **Supplementary Table 1** and **Supplementary Table 2**, respectively. **Figure S12** illustrates the comprehensive lipoprotein subclass data and the intertwined correlation structure in a metabolic network model.

Key results and the fundamental characteristics of lipoprotein subclass particles and metabolism are highlighted in a summary **Figure 8**.

**Figure 8.**
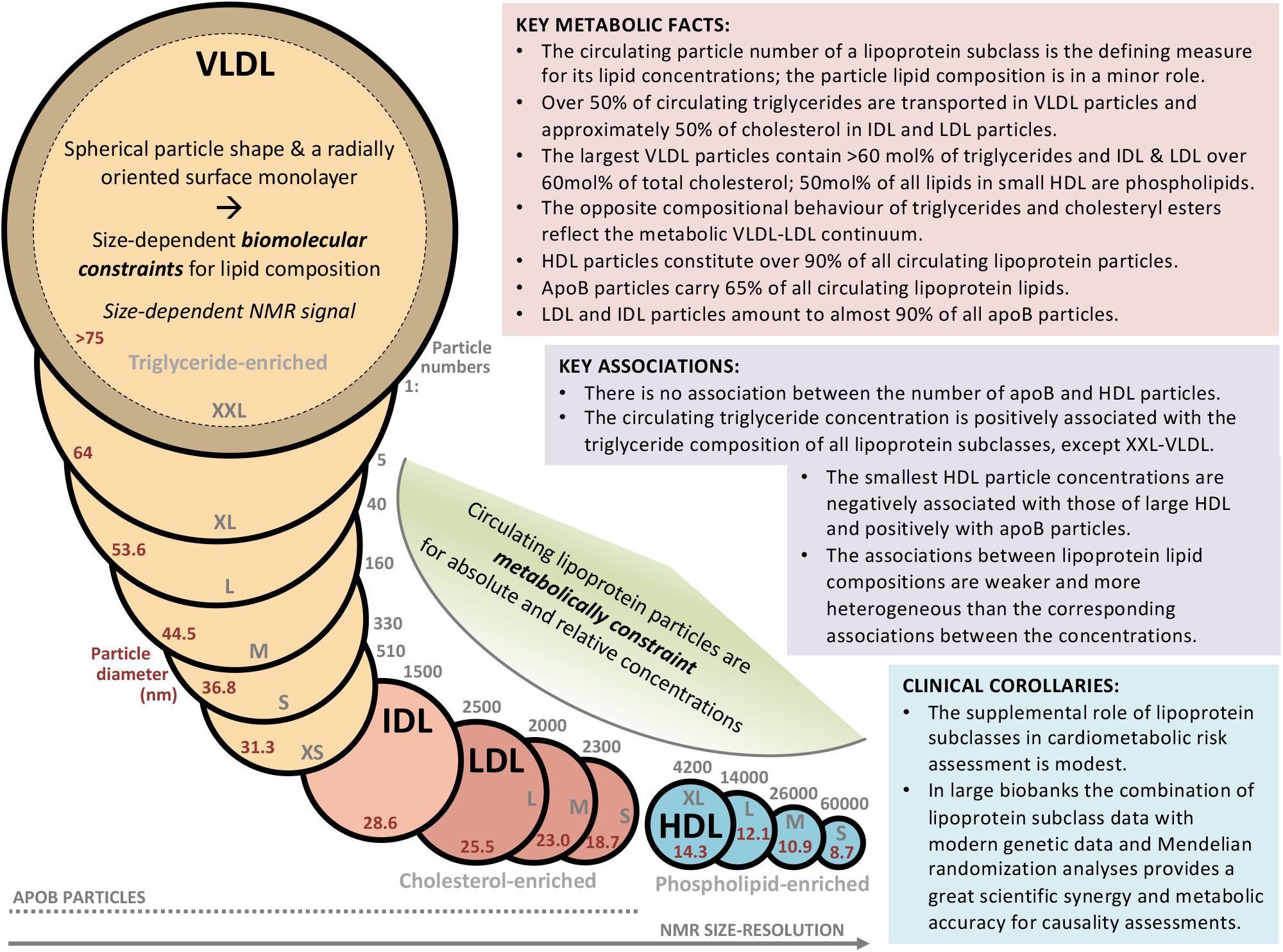
A summary figure encapsulating the key characteristics, associations and metabolic features of the 14 lipoprotein subclasses quantified with the NMR spectroscopy platform.^9,17,19^ The position of the individual lipoprotein subclass particles on the NMR size-resolution axis aims to picture the methodological resolution difference for large and small lipoprotein particles.^6,16^

## Discussion

### Biomolecular and metabolic constraints regarding lipoprotein subclasses

It has been shown earlier that the circulating lipid concentrations as well as the relative lipid compositions for the lipoprotein subclasses with this methodology are very well concordant with those from density-based ultracentrifugation analyses,^4,27^ clinical chemistry^10^ and high-performance liquid chromatography.^17^ The compositional changes in relative to triglycerides and cholesteryl esters are an intrinsic indication of the metabolic continuum in the VLDL-LDL metabolic cascade in the circulation.^13,28,29^ As far as we are aware, the log-linear relationship between circulating lipoprotein subclass particle concentrations and the particle size has not been indicated before. The scale of difference in the subclass particle numbers is huge, *e*.*g*., 60,000 S-HDL particles per one circulating XXL-VLDL particle (**Figure 8**). On average, over 90% of all circulating lipoprotein particles are HDL particles. Even though the particle volumes also differ considerably for different lipoprotein subclasses, the scale is far from the scale in the particle concentrations: the volume of an XXL-VLDL particle is approximately 640 times that of an S-HDL particle. Therefore, the subclass particle concentrations are the main determinants of circulating lipoprotein subclass lipid concentrations and the variation in their lipid compositions is only a minor contributor. However, the view for circulating lipids is quite different from that of the individual particles: only 35% of all lipoprotein lipids are transported in HDL particles.

The key metabolic role of lipoprotein subclass particle concentrations is not unexpected. Within the particles, the size and structural molecular constraints fundamentally define how many non-polar and polar lipid molecules can be accommodated both in the core and surface, respectively. Although the core and surface regions are not rigid, the spherical shape with an oriented monolayer poses strong biophysical constraints within the small number of key lipid classes – triglycerides, phospholipids, free and esterified cholesterol – to be integrated into the particles.^4,5,16^ Thus, functional metabolic issues, *e*.*g*., an increasing dietary fat load, must be solved by increasing the number of chylomicron (exogenously) and VLDL (endogenously) particles to transport the triglycerides; the structural modifications at the particle level would not be able to accommodate changes at that scale due to the biomolecular constraints. This does not mean, however, that the concentration issues would not affect the compositional characteristics (please see below in relation to triglycerides), but it is crucial to appreciate the largely different scale of achievable concentration changes and compositional modifications.

The log-linear relationship between lipoprotein subclass particle concentrations and the particle size suggest an existence of also fundamental metabolic constraints. This means that not only typical particle and lipid concentration ranges are elemental for humans, but also only certain relative lipoprotein subclass particle concentrations are possible in human lipoprotein metabolism. We are not aware of previous discussion directly in relation to this, but we find it implicit in the lipoprotein literature and, for example, via the traditional clinical definitions of abnormalities in lipoprotein metabolism and their drug treatment.^30^ The large-scale data and the high resolution with respect to lipoprotein subclasses only amplifies and clarifies this issue. It is rather logical that a certain (even very high) load of nutritional fats (*i*.*e*., triglycerides) would lead to a corresponding amount of transport particles, exogenously in the form of gut-derived chylomicron/apoB48-containing particles and endogenously via apoB100-containing VLDL particles.^28^ What can metabolically come out of the apolipoprotein B48/B100-containing lipoprotein cascades in the form of remnant, IDL and LDL subclass particles in the circulation, must be in a direct relationship with the secreted amount of chylomicron and/or VLDL particles. The amount of HDL particles partly couples with this cascade though also other sources and metabolic processes for HDL particles and their variation exist.^31^ Notably apolipoprotein B and A-I concentrations (or apolipoprotein B-containing lipoprotein and HDL particle concentrations) only associate very weakly per se. This is explained by the opposite associations between apolipoprotein B and the larger/medium versus the smallest HDL particles. Altogether it is likely that these metabolic constraints are just simple reflections of normal metabolic pathways on how the human body handles triglycerides, cholesteryl esters and other dietary lipids. A key corollary, however, is that when evaluating a risk related to a lipoprotein subclass, it would only be relevant to estimate it within the physiologically realistic concentration area.^21–24^

### Standard lipids, lipoprotein subclasses and their lipid composition

The associations between the standard lipid measures and the subclass concentration measures are intuitive based on how the subclass particle categories are included in the standard lipid measures. For example, HDL cholesterol is the cholesterol carried in all the HDL subclass particles in the circulation and the triglyceride concentration refers to the sum of triglycerides in all the lipoprotein subclass particles. In the case of LDL cholesterol, the situation is clear within the NMR analysis, *i*.*e*., it represents the sum of cholesterol in all LDL subclass particles. Thus, the LDL cholesterol based on the NMR platform is size-specific and does not include cholesterol in any other particles. Generally, however, “LDL” as a concept is far from clear since multiple “LDL” cholesterol assays exist based on remarkably varying methodologies, leading to between-assay heterogeneity in the values of “LDL” cholesterol. The assay-dependent ambiguity can lead to either under-or over-estimation of the true (size-specific) LDL cholesterol. We have recently discussed these issues and their potential implications in detail.^14^ The size-specific accurate definition of LDL and other lipoprotein subclasses is a definite virtue of the NMR-based lipoprotein analytics.

The overall association pattern for the lipoprotein subclass concentration measures is structured and characterised by strong correlations between adjacent apolipoprotein B-containing lipoprotein subclasses with clearly diminishing associations along the metabolic continuum from VLDL to LDL particles. On the contrary, the association pattern for the lipoprotein subclass composition measures is assorted and lacks pronounced patterns. Overall triglyceride and cholesteryl ester compositions tend to associate negatively, particularly within the major lipoprotein fractions VLDL, IDL and LDL, and HDL. A consolidated interpretation of all the associations between the concentration and composition measures is the consistent behaviour of triglycerides. High circulating triglyceride concentrations are reflected by high concentrations of triglyceride-enriched VLDL particles that appear to cause a metabolic spillover effect and result in coherently triglyceride-enriched particles in all lipoprotein subclasses. The smallest HDL subclass behaves similarly to apolipoprotein B-containing subclasses regarding various associations, in contradiction to the other HDL subclasses.

### Large-scale epidemiology in relation to lipoprotein subclass data

This particular NMR platform has been applied in a few large-scale prospective cardiovascular outcome studies with the focus on lipoprotein subclass concentrations.^21–24,32^ The lipoprotein subclass risk profile for cardiovascular events in a European population^21^ and for incident myocardial infarction in a Chinese population^22^ appears highly concordant. The subclass risk profile is similar between incident myocardial infarction and ischemic stroke, but differs considerably for intracerebral haemorrhage for which none of the lipoprotein subclasses appears as a risk factor.^22^ There are also apparent differences in how the lipoprotein subclasses associate with peripheral and coronary artery disease, the associations being weaker for the former.^24^ Applications in other areas, as reviewed by Würtz *et al*. in 2017,^10^ have also almost entirely utilised only lipoprotein subclass concentration data.^10^ Most recent large-scale applications involve studies on, *e*.*g*., oral glucose tolerance test,^33^ healthy diets and risk of cardiovascular disease,^34^ and exome sequencing for rare variants in cardiometabolic traits.^35^

### Large-scale epidemiology in relation to compositional data on lipoprotein subclasses

Only a few large-scale epidemiological studies have exploited the compositional lipoprotein subclass data from this NMR platform or in general. Laakso and co-workers studied lipoprotein subclass profiles in individuals with varying degrees of glucose tolerance in a population-based study of over 9,000 Finnish men.^20^ Concomitantly to increased serum concentrations of essentially all apolipoprotein B-containing lipoprotein particles as well as the smallest HDL particles, hyperglycaemia and insulin resistance were associated with triglyceride-enrichment in various subclass particles, although these differences were related more to hypertriglyceridaemia than to hyperglycaemia.^20^ Barrios et al.^36^ noted that in both diabetic and non-diabetic individuals the triglyceride composition of various apolipoprotein B-containing and some HDL particles were rather coherently inversely associated with kidney function and microvascular complications of diabetes. A recent study examined observational associations of all lipoprotein subclass measures with risk of incident CHD in three population-based cohorts totalling 616 incident cases and 13,564 controls during 8-year follow-up.^13^ This study was the first large-scale study to assess the independent role of lipoprotein subclass composition in CHD risk assessment with the finding that a higher triglyceride composition within HDL subclasses was associated with higher risk of CHD, independently of total cholesterol and triglycerides. The largest hazard ratio for the triglyceride-enrichment in medium HDL subclass particles was of a similar magnitude as that for LDL cholesterol and apolipoprotein B, *i*.*e*., around 1.3. However, the findings were interpreted to reflect the combined effects of circulating HDL and apolipoprotein B-containing particles, maybe in connection to CETP function and the circulating amount of total triglycerides, not an intrinsic indication of an independent role of HDL particle lipid composition in CHD.^13^

### Drug-target Mendelian randomization

The lipoprotein subclass data have recently been actively utilised in drug-target Mendelian randomization analyses.^12,13,15,37,38^ In these studies specific genetic instruments are used as a proxy for the pharmacological on-target effects of proteins to understand and quantify the involvement of a protein in disease.^37–39^ The detailed size-resolution provided by the NMR-based lipoprotein data has been advantageous in the detailed appreciation of the intricate details and differences in various cholesterol lowering drugs.^12,15,37^ The comparison between the results from actual randomised controlled drug trials and the drug-target Mendelian randomization is compelling, as recently demonstrated in a prospective pravastatin trial together with comparisons to the metabolic effects of genetic inhibition of HMG-CoA reductase (statins) and proprotein convertase subtilisin/kexin type 9 (PCSK9).^12^ A drug-target Mendelian randomization study on CETP inhibition revealed that conventional composite lipid assays (*e*.*g*., “LDL cholesterol”)^14^ may mask important heterogeneous effects on lipoprotein profiles.^13,40^ The differences between the observed drug effects on traditional “LDL cholesterol” and, both analytically and metabolically, more specific apolipoprotein B-containing lipoprotein subclasses are calling attention to the need for metabolic precision in the measurements of lipoprotein data and in assessing the role of lipoprotein metabolism in cardiovascular disease in relation to existing and novel lipid-altering therapies. A particularly problematic issue would be to use a nonspecific marker of lipids to derive an expected effect of a drug with risk of disease.^13,14^ The combination of NMR-based lipoprotein subclass data with drug-target Mendelian randomization analyses has shown a great synergy for drug development in the current era of biobanks and big data. However, it should be kept in mind that the use of drug-target Mendelian randomization analysis can principally provide information only on pharmacological on-target effects (of a class of drugs) and does not allow for assessing potential off-target effects (of individual drugs).^41^

### Lipoprotein subclass measures as risk factors

The high size-resolution offered by the NMR-based lipoprotein profiling has resulted in a detailed understanding of how the various stages and remnant particles in the apolipoprotein B-containing lipoprotein cascade relate to cardiometabolic issues and outcomes.^10,21,22,29,42^ However, from the predictive risk factor perspective the additional information provided by the lipoprotein subclass particle concentrations in common cardiovascular disease outcomes appears inconsequential, particularly keeping in mind the current recognition of the key role of apolipoprotein B in the development of atherosclerosis and CHD.^13,21,43–45^ The potential role of lipoprotein particle compositions (as independent of concentrations) has occasionally been discussed in the literature since the advent of sequential preparative ultracentrifugation for lipoprotein analytics but the tediousness and cost of the process have precluded large-scale studies.^2,13^ The clinically most utilised NMR-based lipoprotein analytics unfortunately does not provide compositional lipoprotein subclass data.^7,46^ Thus, the first large-scale study on the independent role of lipoprotein particle compositions on CHD was published only very recently and based on the NMR methodology on focus here.^13^ However, as discussed above, the most pronounced findings regarding the increased risk in relation to triglyceride-enrichment of HDL particles, might just be a reflection of high circulating triglycerides and not an intrinsic indication of an independent role of HDL particle lipid composition in CHD.^13^ Nonetheless, in type 1 diabetes triglyceride-cholesterol imbalance has been found as a general lipoprotein characteristic indicating high vascular disease risk.^47^

## Limitations

The separation of lipoprotein subclasses in NMR is solely based on particle size. Fundamentally the resolution via NMR chemical shifts is the best for the smallest particles and the weakest for the largest particles (**Figure 8**).^16^ In postprandial conditions the large chylomicron particles will be detected in the largest particle category XXL-VLDL without the possibility to directly resolve VLDL and chylomicron particles. In addition, if smaller apoB48-containing particles are secreted by the gut to the bloodstream,^28^ they are detected purely based on their particle size without the possibility to further identification. This limitation applies also to all potential remnant particles; the detection is only based on the particle size and further metabolic identification, for example, between a VLDL or a chylomicron remnant, is not possible, *i*.*e*., all will be summed up together according to their particle size.

The highly correlated nature of lipoprotein subclass concentration measures, particularly for adjacent subclasses, is sometimes identified as a limitation or a problem. However, this is a direct representation of human lipoprotein metabolism; it is a continuum, particularly for the apolipoprotein B-containing lipoprotein subclasses. Thus, the adjacent subclass particles, both in concentrations and composition, are inevitably very similar with highly correlated attributes. Particularly at high size-resolution of the particles, any other correlation behaviour between measures for adjacent or closely related lipoprotein subclasses would most likely be an indication of methodological hurdles. Nevertheless, the high correlations make part of the information redundant from a strictly statistical perspective but on the other hand, in the case of continuous metabolic processes, high resolution allows for intricate interpretations and findings as already demonstrated in various publications.^10,12,13,15,29,37^

## Data Availability

The datasets used in the current study are available from the cohorts through application process for researchers who meet the criteria for access to confidential data: https://www.oulu.fi/nfbc/.

## Conclusions

The various analyses presented here reveal fundamental characteristics and differences in lipoprotein subclass concentration and composition data. We would urge clear nomenclature in relation to these measures and noting consistently when particle concentrations (mol/L) are meant and with the lipid data, for example, if a triglyceride measure would refer to a circulating concentration in a particular subclass of particles (mmol/L) or to a relative composition in a particular particle subclass (mol% of total lipids). The overall spherical structure of these particles, with an oriented surface monolayer, results in strong size-dependent biomolecular constraints for their lipid compositions. In addition, the metabolic fundaments in humans control the possible circulating lipoprotein subclass particle and lipid concentrations both in absolute and relative terms. Comprehensive data on lipoprotein subclass particles in large-scale studies provide opportunities for detailed understanding of lipoprotein metabolism in health and disease. Combining these data with modern genetics in exceedingly extensive studies as the UK Biobank, will facilitate grasping what features might represent causal, and thus, potentially treatable and druggable, metabolic risk factors.

## Supplementary material

Supplementary material is available at the Journal online.

## Funding

MAK is supported by a research grant from the Sigrid Juselius Foundation, Finland. PO is supported by the Emil Aaltonen Foundation. The Northern Finland Birth Cohorts have received funding from the Academy of Finland, Novo Nordisk Foundation, and EU.

## Disclosure

The authors declare that there is no conflict of interest.

## Notes

### Competing Interest Statement

The authors have declared no competing interest.

### Author Declarations

The studies comply with the Declaration of Helsinki, were approved by the local ethics committee (Northern Ostrobothnia Hospital District, Finland) and written informed consents were obtained from each participant.

