## Supplement for "Apt interpretation of comprehensive lipoprotein data in large-scale epidemiology – disclosure of fundamental structural and metabolic relationships"

### Supplement content

**Supplement Note**

**Study populations**

**Supplement Tables**

**Table S1.** Spearman's rank correlations (adjusted for sex) for the lipoprotein subclass concentration (98), composition (70) and standard lipid (4) measures as well as apolipoprotein B and A-I for the 5,651 participants in NFBC66 (Figure S10).

**Table S2.** Spearman's rank correlations (adjusted for sex) for the lipoprotein subclass concentration (98), composition (70) and standard lipid (4) measures as well as apolipoprotein B and A-I for the 5,605 participants in NFBC86 (Figure S11).

**Supplement Figures**

**Figure S1.** The relative lipid compositions of the 14 lipoprotein subclasses for the 5,605 participants in NFBC86.

**Figure S2.** The circulatory and compositional characteristics of lipoprotein subclasses for the 5,605 participants in NFBC86.

**Figure S3. A.** The distributions of particle concentrations for each lipoprotein subclass for the 5,605 participants in NFBC86. **B.** Various proportions of lipoprotein particles and lipids for the 5,605 participants in NFBC86.

**Figure S4.** The log-linear relationship between the circulating lipoprotein subclass particle concentration and the particle diameter (5,605 participants in NFBC86).

**Figure S5.** The absolute circulating lipid concentrations for each lipoprotein subclass and the corresponding summary measures for the apolipoprotein B-containing lipoprotein particles and HDL particles for the 5,605 participants in NFBC86.

**Figure S6.** For the 5,605 participants in NFBC86: **A.** The associations between traditional lipoprotein lipid measures. **B.** The associations between traditional lipoprotein lipid measures and lipoprotein subclass particle and triglyceride concentrations. **C.** The associations between traditional lipoprotein lipid measures and lipoprotein subclass lipid composition for triglycerides and cholesteryl esters. **D.** The associations between lipoprotein subclass particle and triglyceride concentrations and lipoprotein subclass lipid composition for triglycerides and cholesteryl esters.

**Figure S7.** Scatter plots for the 5,651 participants in NFBC66: **A.** Apolipoprotein B concentration vs apolipoprotein B particle concentration. **B.** Apolipoprotein A-I concentration vs HDL particle concentration. **C.** Apolipoprotein B concentration vs apolipoprotein A-I concentration. **D.** Apolipoprotein B particle concentration vs HDL particle concentration.

**Figure S8.** Scatter plots for the 5,651 participants in NFBC66: **A.** Apolipoprotein B particle concentration vs XL-HDL particle concentration. **B.** Apolipoprotein B particle concentration vs L-HDL particle concentration. **C.** Apolipoprotein B particle concentration vs M-HDL particle concentration. **D.** Apolipoprotein B particle concentration vs S-HDL particle concentration.

**Figure S9.** For the 5,605 participants in NFBC86: **A.** The associations between lipoprotein subclass particle and triglyceride concentrations. **B.** The associations between lipoprotein subclass lipid composition for triglycerides and cholesteryl esters.

**Figure S10.** A heatmap of Spearman's rank correlations (adjusted for sex) for the lipoprotein subclass concentration (98), composition (70) and standard lipid (4) measures as well as apolipoprotein B and A-I for the 5,651 participants in NFBC66 (Table S1).

**Figure S11.** A heatmap of Spearman's rank correlations (adjusted for sex) for the lipoprotein subclass concentration (98), composition (70) and standard lipid (4) measures as well as apolipoprotein B and A-I for the 5,605 participants in NFBC86 (Table S2).

**Figure S12.** A Spearman's rank correlation network between lipoprotein subclasses for the 5,651 participants in NFBC66.

### Supplement Note

#### Study populations

The Northern Finland Birth Cohort (NFBC) studies are two longitudinal birth cohorts established to study factors affecting preterm birth and consequent morbidity in the two northernmost provinces of Finland, Oulu and Lapland. The NFBC66 includes 12,058 live births (12,231 children) covering 96% of all eligible births in this region during January – December 1966.[1] The participants were followed-up at the age of 1, 14, 31 and 46 years. Data collection conducted in 2012 at their age of 46 years, including clinical examination and fasting serum sampling and NMR spectroscopy, was available for 5,651 participants and these data were used for the main analyses in this work. Two decades later, a second cohort of 9,432 births was collected (NFBC86) which covered 99% of all the deliveries taking place in the target regions during July 1985 – June 1986.[2] Data collection in 2001–2002 including clinical examination, fasting serum samples and NMR data at the age of 15–16 was available for 5,605 adolescents and was used as a replication in this work. The studies comply with the Declaration of Helsinki, were approved by the local ethics committee (Northern Ostrobothnia Hospital District, Finland) and written informed consents were obtained from each participant.

### Supplement Figures

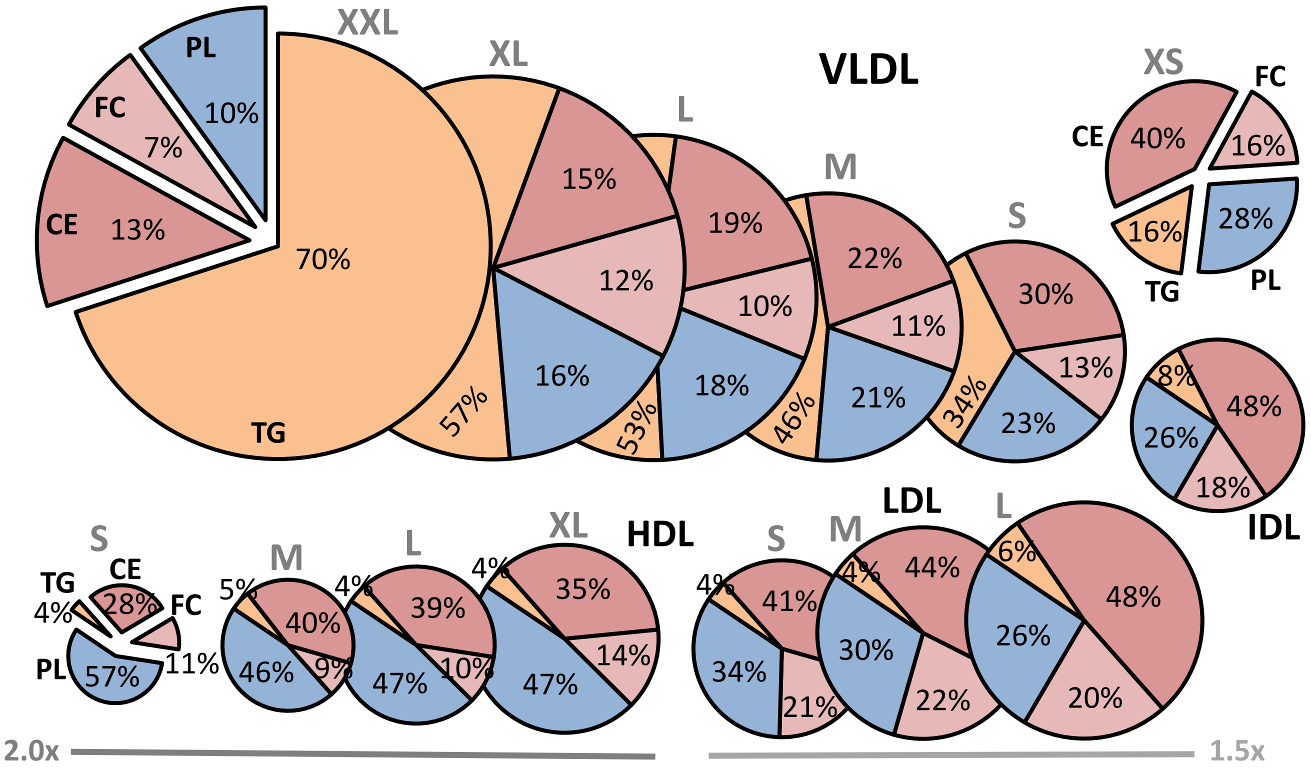


**Figure S1**

The relative lipid compositions of the 14 lipoprotein subclasses. The NMR platform resolution is 14 lipoprotein subclasses, defined by their particle size as follows: six VLDL particle categories; XXL-VLDL (with average particle diameter >75 nm), XL-VLDL (64 nm), L-VLDL (53.6 nm), M-VLDL (44.5 nm), S-VLDL (36.8 nm), and XS-VLDL (31.3 nm); IDL (28.6 nm), L-LDL (25.5 nm), M-LDL (23.0 nm), and S-LDL (18.7 nm); and XL-HDL (14.3 nm), L-HDL (12.1 nm), M-HDL (10.9 nm) and S-HDL (8.7 nm). Note that the size of LDL and HDL particles in the figure is multiplied by 1.5 and 2.0, respectively. The data are mean values for 5,605 participants in NFBC86. VLDL, very-low-density lipoprotein; IDL, intermediate-density lipoprotein; LDL, low-density lipoprotein; HDL, high-density lipoprotein; XXL, extremely large; XL, very large; L, large; M, medium; S, small; XS, very small; TG, triglycerides; PL, phospholipids; CE, cholesteryl esters; FC, free cholesterol.

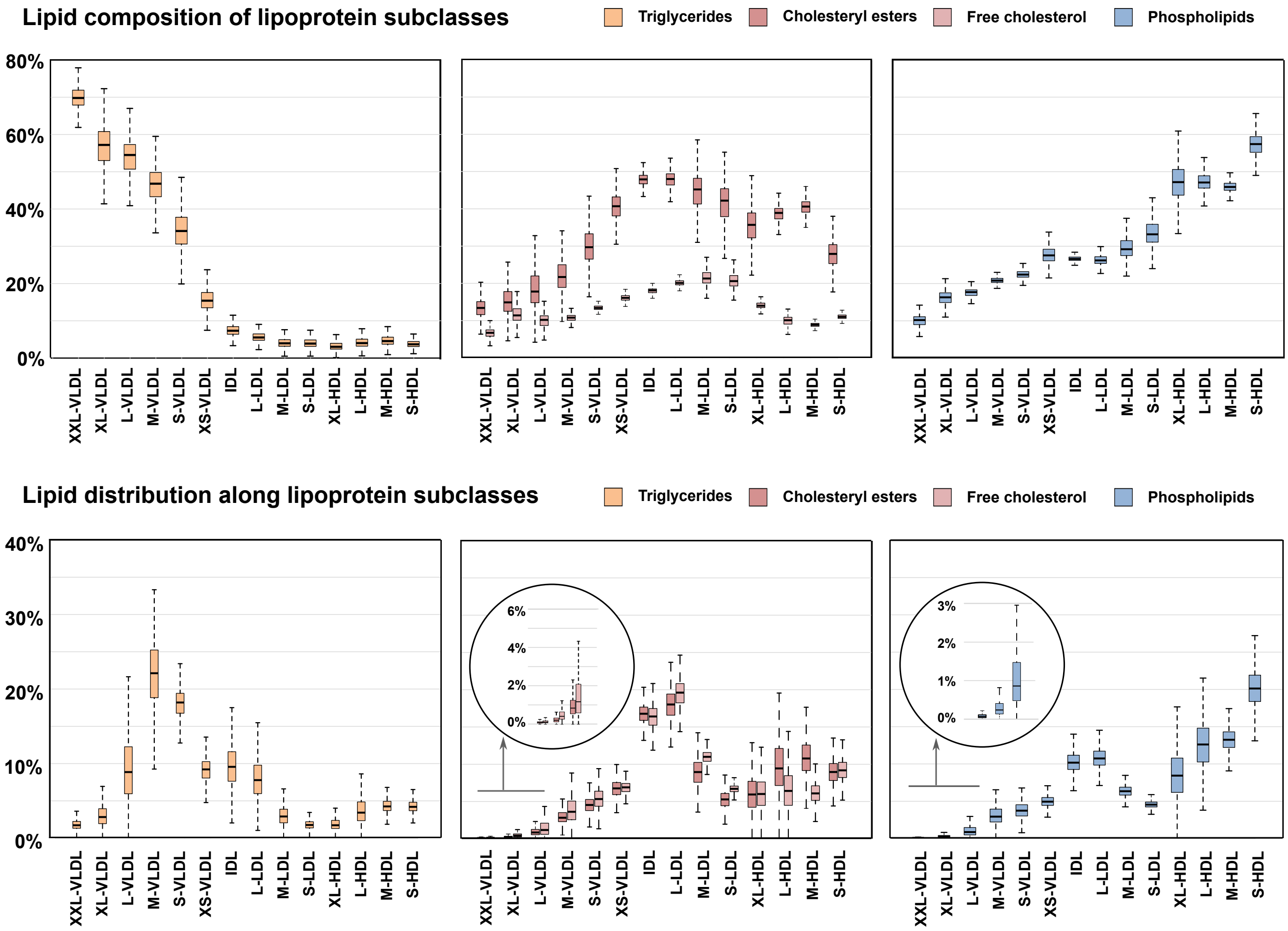


### Figure S2

The circulatory and compositional characteristics of lipoprotein subclasses. The lipid distribution along the lipoprotein subclasses illustrates how a certain lipid class is distributed in the blood stream among all the lipoprotein subclasses (i.e., the sum of all percentages in each inset is 100%). The composition of lipoprotein subclasses depicts what are the relative lipid contents in each subclass particle category (i.e., the sum of the percentages of all the lipid classes for each subclass is 100%). The data are from the NFBC86 including 5,605 participants; each box plot shows the median within the interquartile range (IQR) and the minimum (Q1 - 1.5*IQR) and maximum (Q3 + 1.5*IQR) values with potential outliers. The abbreviations are as explained in the caption for Figure S1.

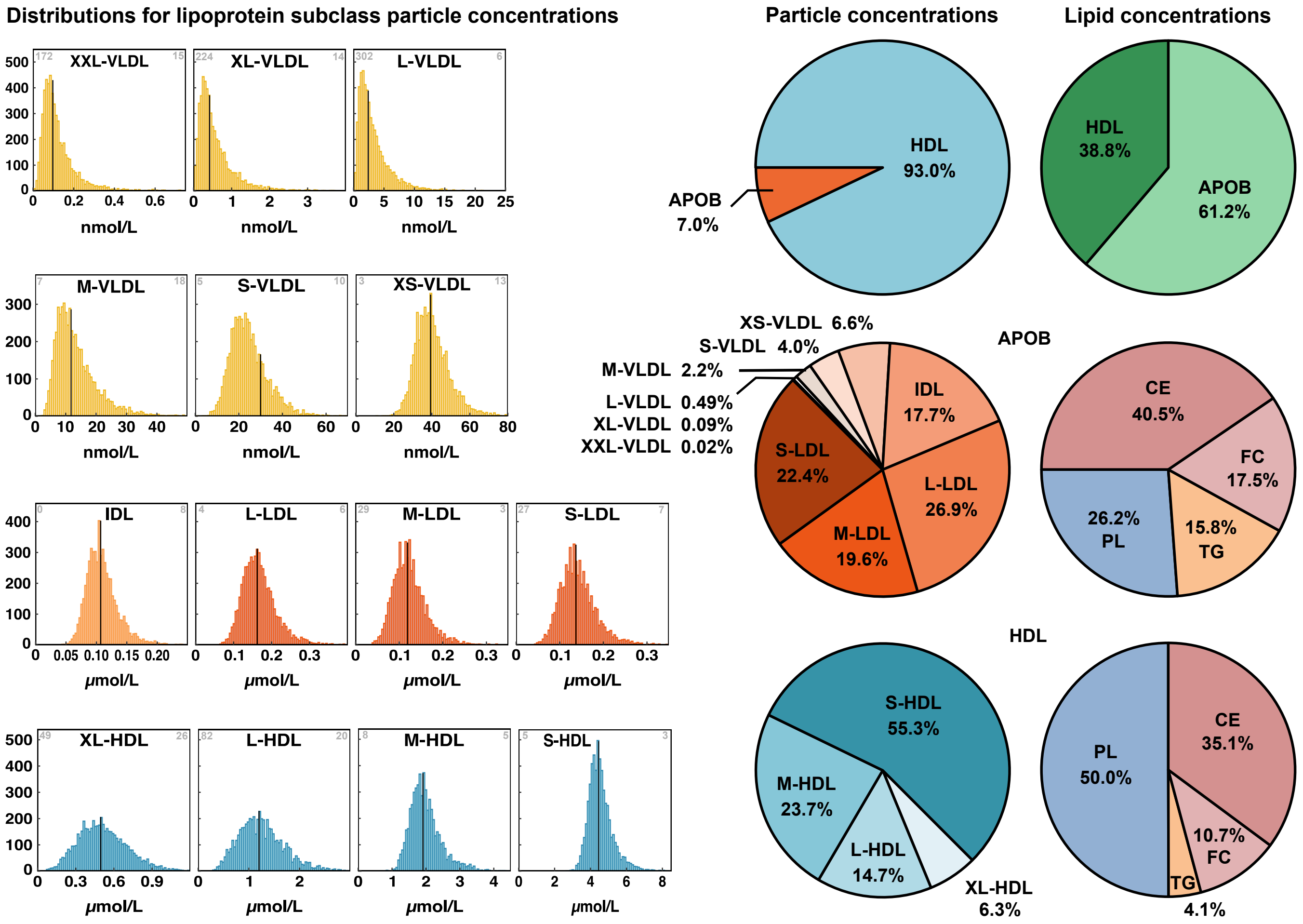


### Figure S3

The distributions of particle concentrations for each lipoprotein subclass. The data are from the NFBC86 including 5,605 participants. The grey number in the upper-left corner identifies the number of samples for which the particle concentration is zero and in the upper-right corner for how many high concentration values were cut off from the drawn distribution. Note that the concentration scale is nmol for the VLDL, IDL and LDL particles and µmol for the HDL particles. The black vertical lines denote the median concentration values. Various proportions of lipoprotein particles and lipids are shown in the pie charts as mean values for the 5,605 participants in NFBC86. The abbreviations are as explained in the caption for Figure S1.

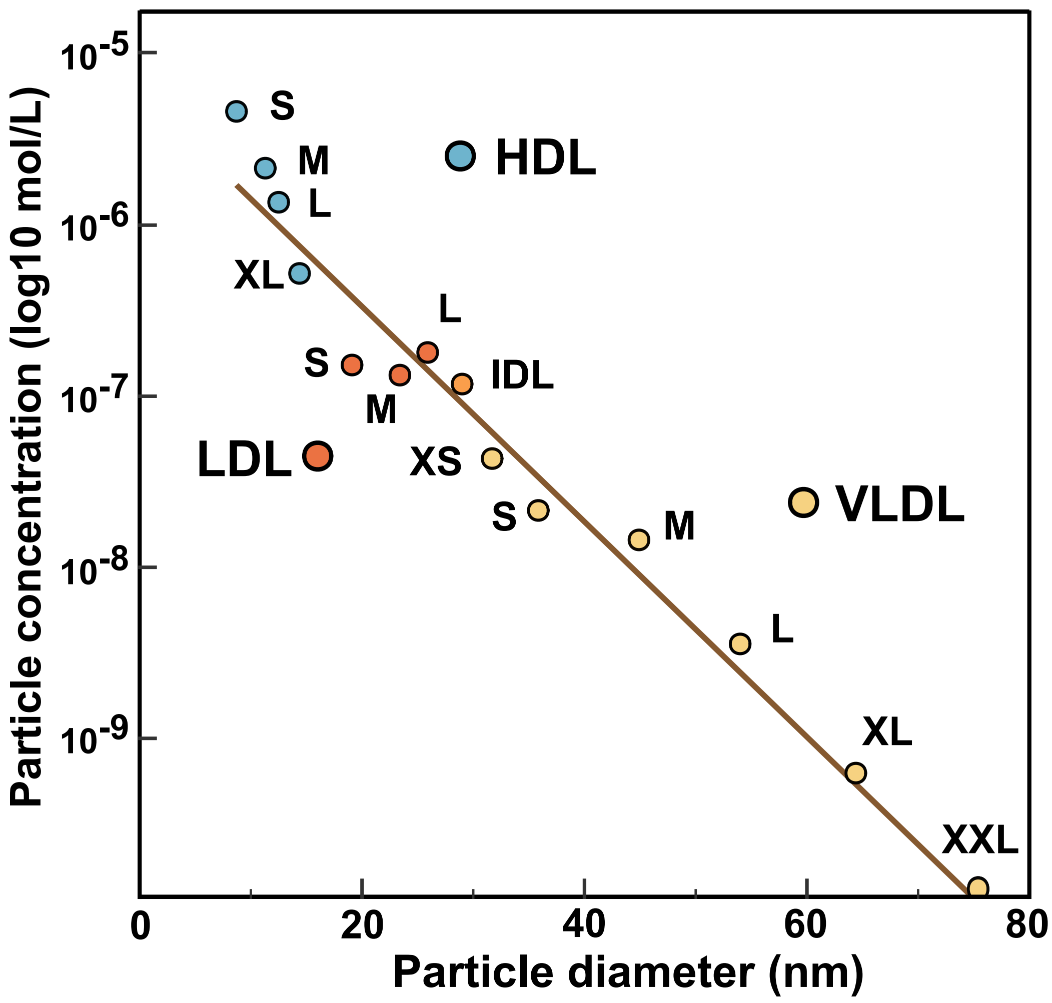


**Figure S4**

The log-linear relationship between the circulating lipoprotein subclass particle concentration and the particle diameter. The data and abbreviations are as explained in the caption for Figure S1.

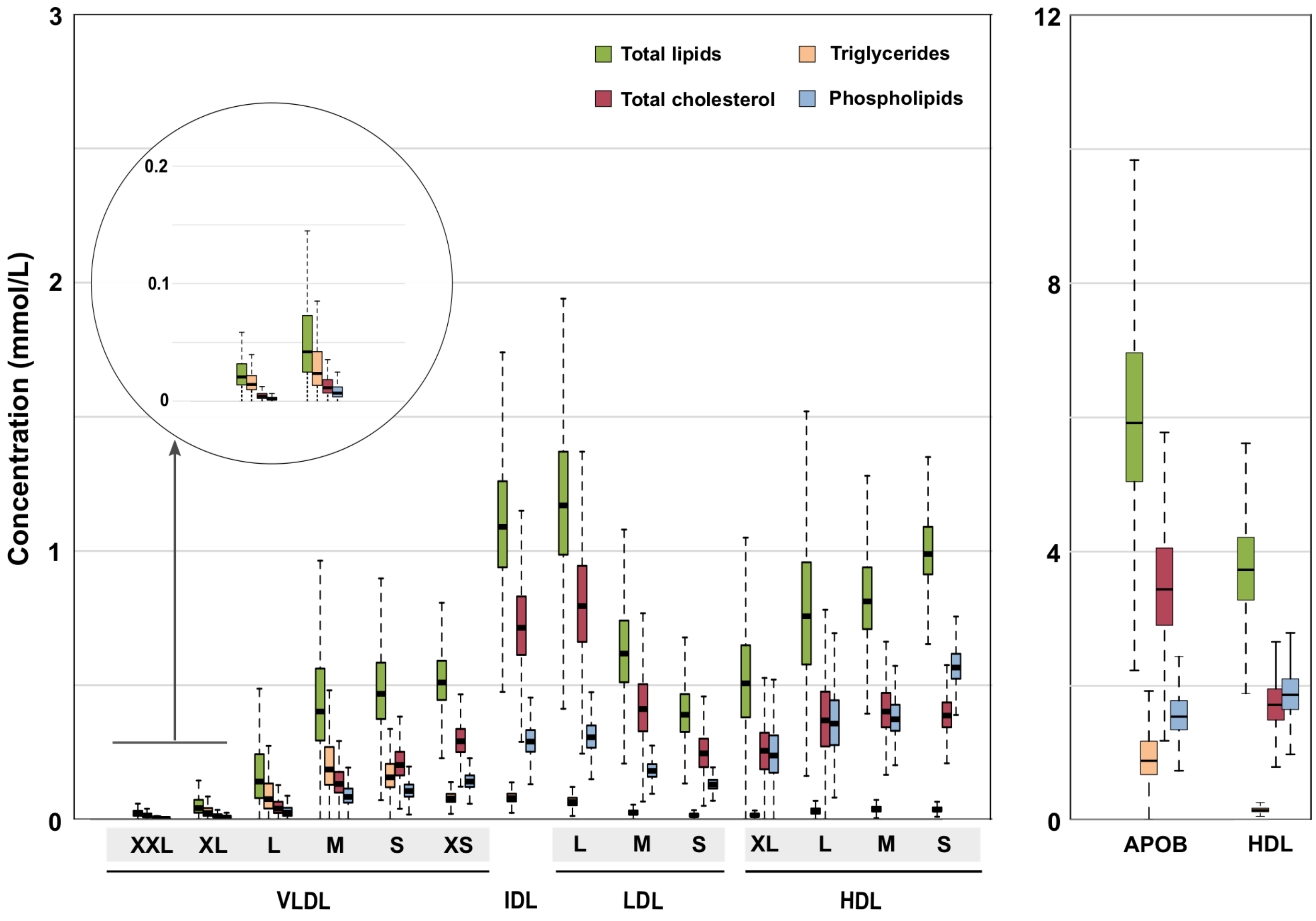


**Figure S5**

The absolute circulating lipid concentrations for each lipoprotein subclass and the corresponding summary measures for the apolipoprotein B-containing lipoprotein particles and HDL particles. The data and abbreviations are as explained in the captions for Figure S1 and S2.

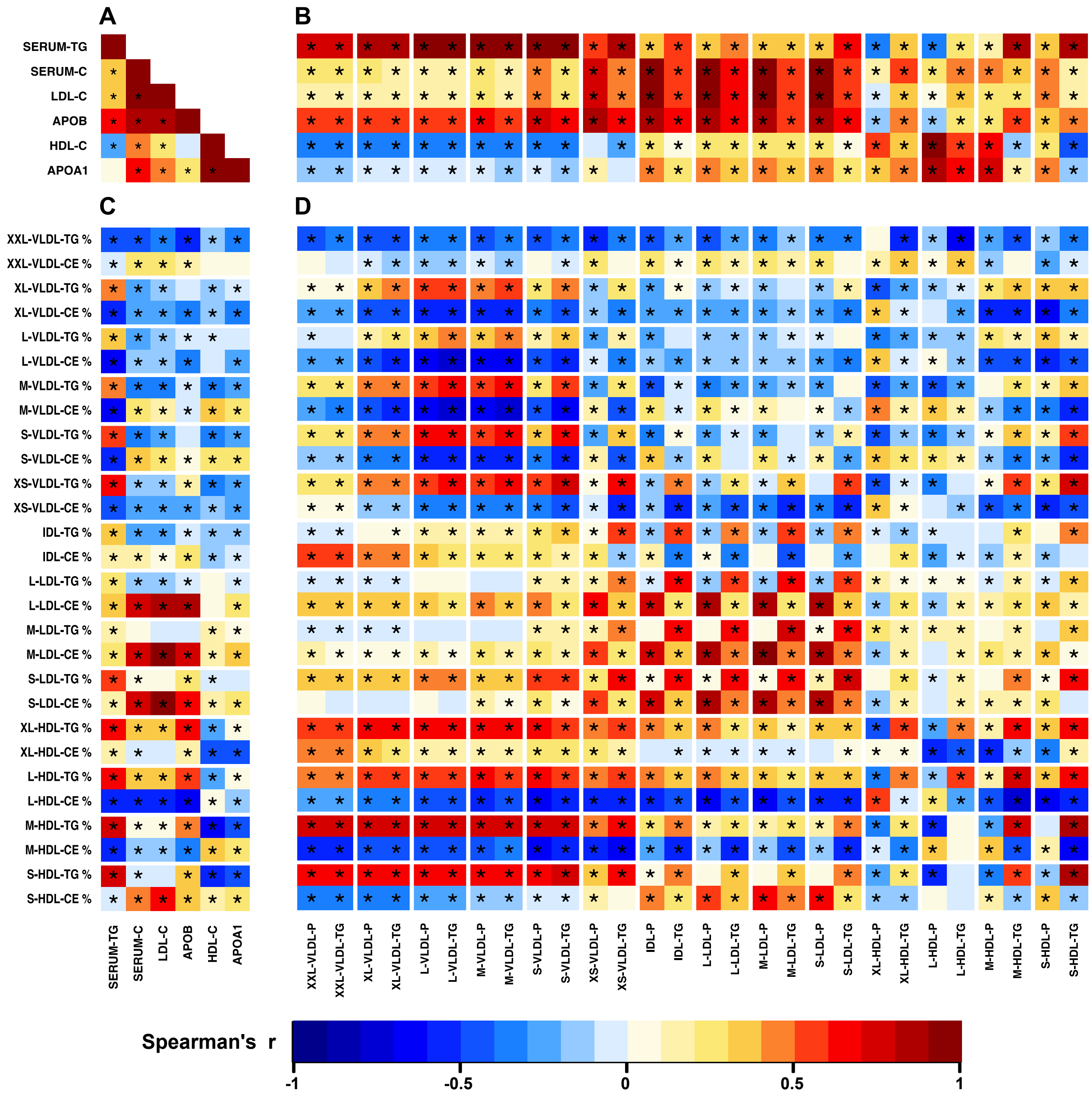


### Figure S6

The associations between traditional lipoprotein lipid measures (A), between traditional lipoprotein lipid measures and lipoprotein subclass particle and triglyceride concentrations (B), between traditional lipoprotein lipid measures and lipoprotein subclass lipid composition for triglycerides and cholesteryl esters (C), and between lipoprotein subclass particle and triglyceride concentrations and lipoprotein subclass lipid composition for triglycerides and cholesteryl esters (D). The colour key refers to Spearman's rank correlation coefficients the NFBC86 including 5,605 participants (adjusted for sex). Twenty-three principal components explained >99% of variation in the 174 lipoprotein measures (98 concentrations and 70 compositions for the lipoprotein subclasses, 4 traditional lipid measures and apolipoprotein A-I and B). Thus, a P-value threshold of 0.002 (*i.e.*, 0.05/25) was used to denote evidence in favour of an association (marked * in the maps). The %-sign refers to the compositional measures (i.e., the percentage of a lipid class concentration of the total lipid concentration in a particular lipoprotein subclass). Serum-TG, total circulating triglyceride concentration; Serum-C, total circulating cholesterol concentration; LDL-C, LDL cholesterol; APOB, apolipoprotein B; HDL-C, HDL cholesterol; APOA-I, apolipoprotein A-I. Other abbreviations are as explained in the caption for Figure S1.

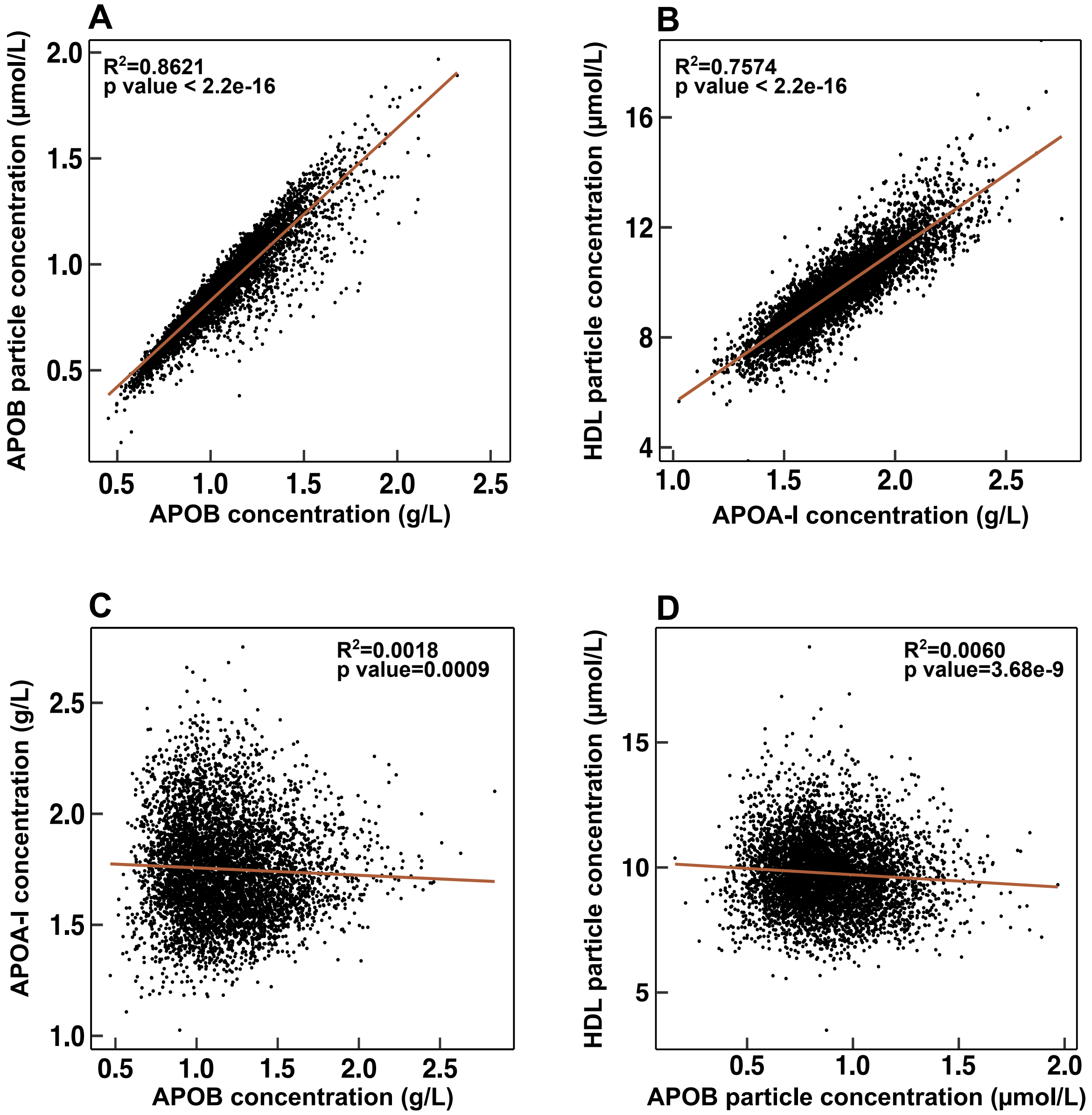


**Figure S7**

Scatter plots for the 5,651 participants in NFBC66: **A.** Apolipoprotein B concentration (g/L) vs apolipoprotein B particle concentration (µmol/L). **B.** Apolipoprotein A-I concentration (g/L) vs HDL particle concentration (µmol/L). **C.** Apolipoprotein B concentration (g/L) vs apolipoprotein A-I concentration (g/L). **D.** Apolipoprotein B particle concentration (µmol/L) vs HDL particle concentration (µmol/L).

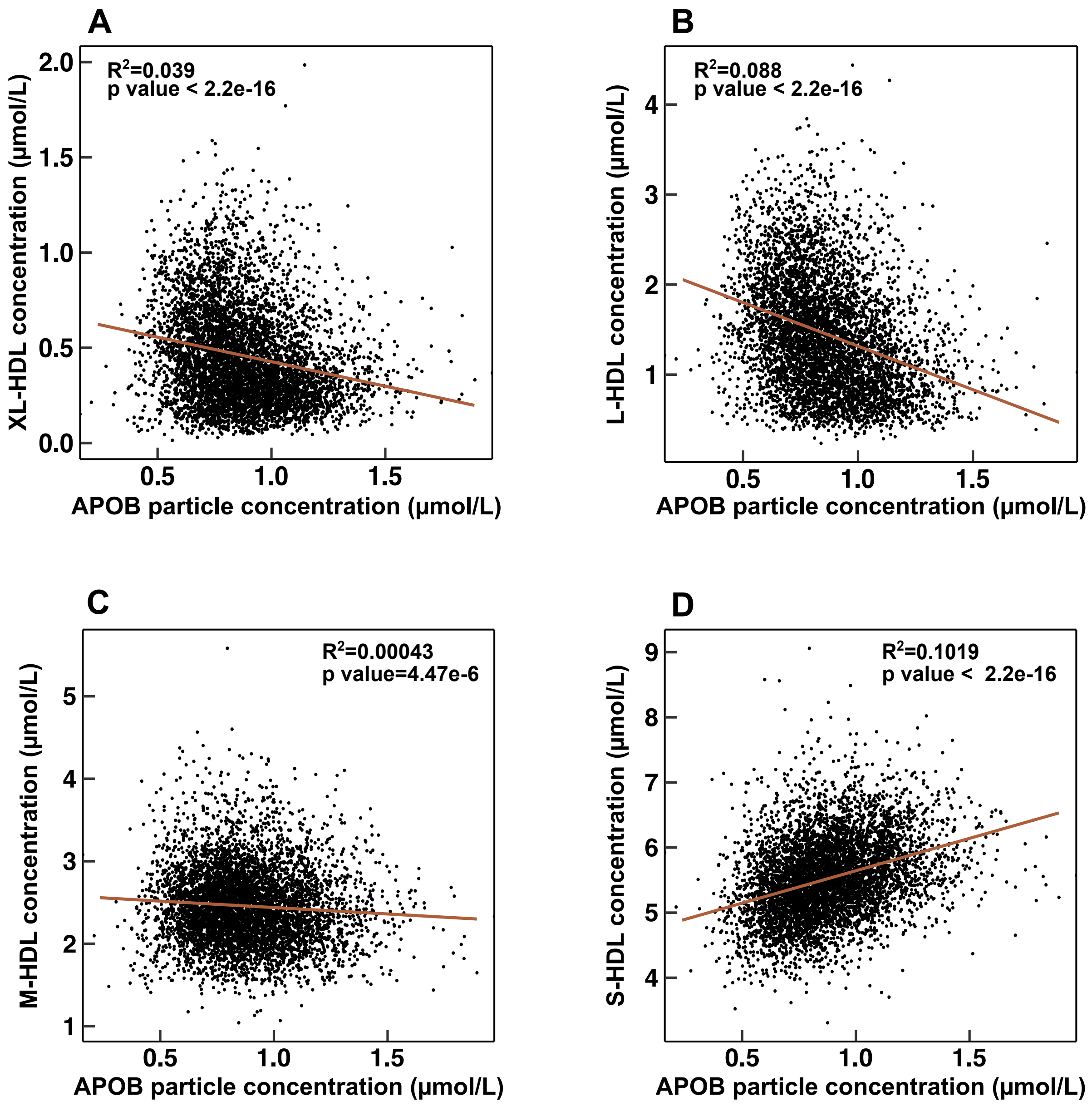


**Figure S8.**

Scatter plots for the 5,651 participants in NFBC66: **A.** Apolipoprotein B particle concentration vs XL-HDL particle concentration. **B.** Apolipoprotein B particle concentration vs L-HDL particle concentration. **C.** Apolipoprotein B particle concentration vs M-HDL particle concentration. **D.** Apolipoprotein B particle concentration vs S-HDL particle concentration.

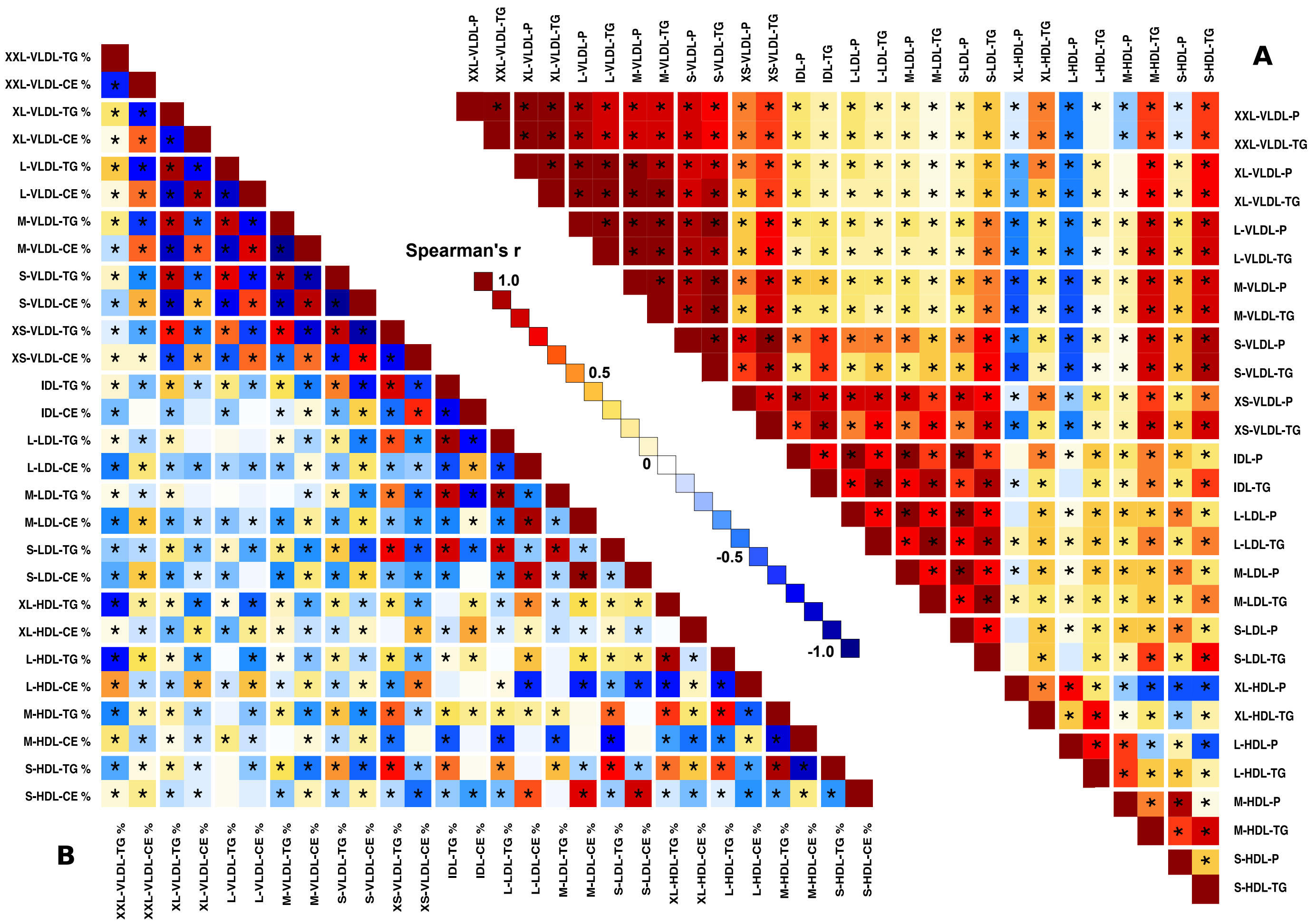


**Figure S9**

The associations between lipoprotein subclass particle and triglyceride concentrations (A) and between lipoprotein subclass lipid composition for triglycerides and cholesteryl esters (B). The data and abbreviations are as explained in the caption for Figure S6.

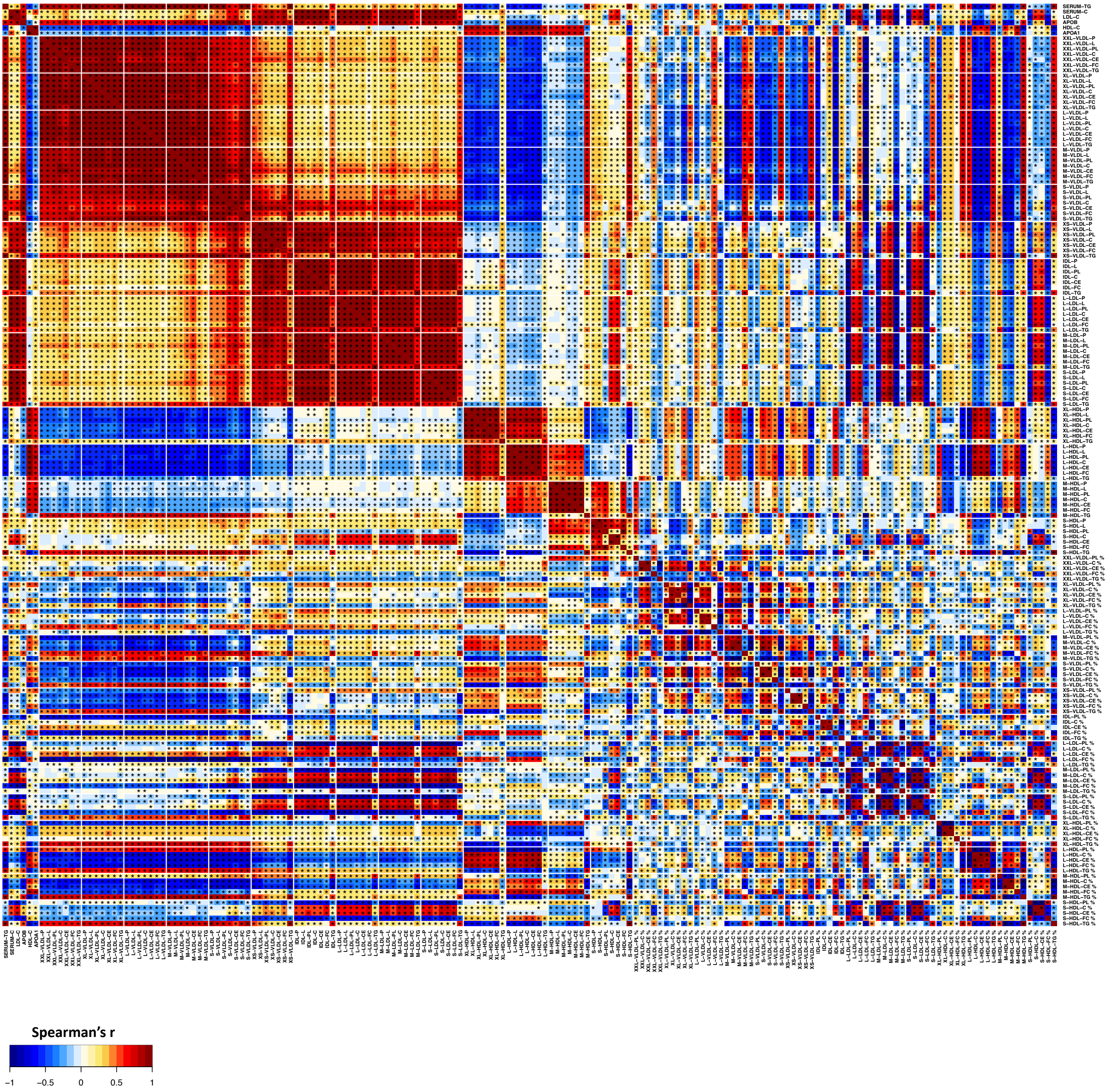


**Figure S10.**

A heatmap of Spearman's rank correlations (adjusted for sex) for the lipoprotein subclass concentration (98), composition (70) and standard lipid (4) measures as well as apolipoprotein B and A-I for the 5,651 participants in NFBC66 (Table S1). The abbreviations are as explained in the caption for Figure S6. L as part of the abbreviation for a subclass measure refers to total lipid concentration and P for total particle concentration.

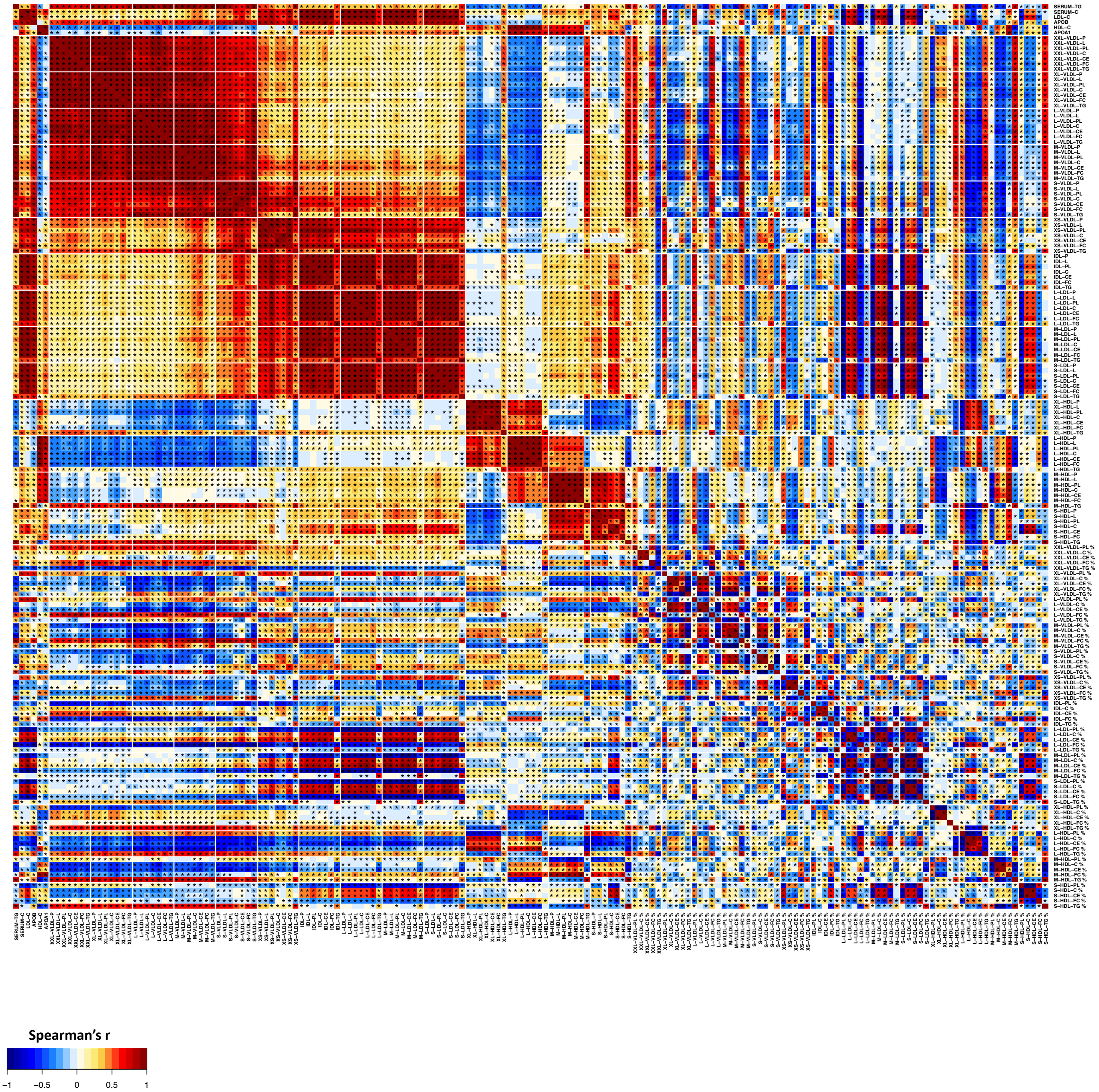


**Figure S11.** A heatmap of Spearman's rank correlations (adjusted for sex) for the lipoprotein subclass concentration (98), composition (70) and standard lipid (4) measures as well as apolipoprotein B and A-I for the 5,605 participants in NFBC86 (Table S2). The abbreviations are as explained in the caption for Figure S6. L as part of the abbreviation for a subclass measure refers to total lipid concentration and P for total particle concentration.

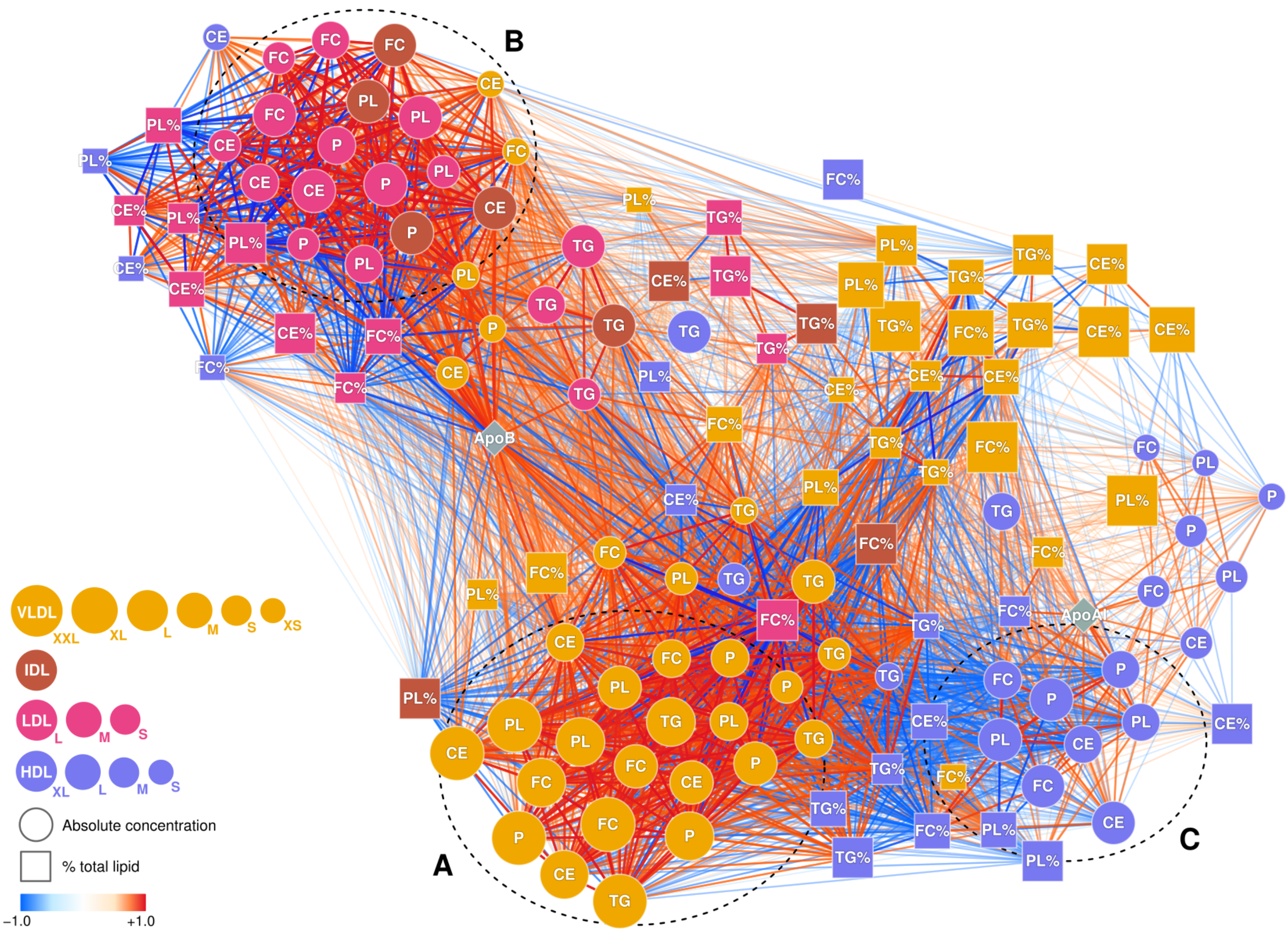


**Figure S12.** A Spearman's rank correlation network between lipoprotein subclasses for the 5,651 participants in NFBC66. Any edges within a subclass were excluded. Derived measures including total lipid and total cholesterol were also excluded. The visualization was created by a modified version of the Fruchterman-Reingold force-directed node-positioning algorithm. Three densely connected groups of variables were observed: **A)** Lipid and particle concentrations of VLDL subclasses, **B)** Lipid and particle concentrations of IDL and LDL subclasses, **C)** Lipid and particle concentrations of the three largest HDL subclasses. The VLDL group (**A**) is positively connected to the IDL/LDL group (**B**) via the apolipoprotein B node in the middle, reflecting the apolipoprotein B particle cascade and metabolic continuum from the triglyceride-rich VLDL subclasses (**A**) to the cholesterol-rich IDL and LDL particles (**B**) in the circulation. The HDL subclass group (**C**) is inversely correlated with the VLDL subclass group (**A**) as an overall reflection of the population level negative correlation between circulating triglycerides and HDL cholesterol. Apolipoprotein A-I node is widely connected among all the HDL subclasses but does not constitute a strong metabolic linkage as apolipoprotein B. The cholesterol and phospholipid compositions of the smallest HDL subclass are strongly connected to the IDL/LDL group (**B**) while the triglyceride composition and concentration are attached to both VLDL subclasses (**A**) and the other HDL subclasses (**C**). Triglyceride concentrations and subclass compositions are strongly positively linked throughout the entire network and among almost all the subclasses demonstrating wide-ranging spillover of circulating triglycerides.
